# Linking Outdoor Air Temperature and SARS-CoV-2 Transmission in the US Using a Two Parameter Transmission Model

**DOI:** 10.1101/2020.07.20.20158238

**Authors:** Ty Newell

## Abstract

Outdoor temperature lower than 50F and greater than 70F is shown to nearly double the transmission efficiency of the SARS-CoV-2 virus. Outdoor temperature is an important factor behind the current surge in US Covid-19 cases. Correlation of northern state infection data and outdoor temperatures is used to identify the change in disease transmission efficiency as northern states passed through the lower temperature bound (50F) in spring, and more recently transitioned to temperatures above the higher bound (70F). At current disease transmission efficiency levels, social distancing must be increased above a UMD Social Distance Index (SDI) level of 36 to stop the accelerated increase of daily infection cases. At current disease transmission efficiency (G=0.19) and SDI of 33, the US will approach 150,000 infections per day in September before declining as average US temperature falls below 70F.

A primary reason for enhanced disease transmission below 50F and above 70F is attributed to inadequate indoor ventilation. Swing season occurs when outdoor temperatures are between 50F and 70F, and is the time of year when homes and buildings are opened to the outdoors. Increased fresh air ventilation (greater than 40cfm per person), improved air filtration (MERV11 and greater filters), and UVGI (Ultraviolet Germicidal Irradiation, 0.02W_UV_ per cfm airflow) coupled with wearing face masks, 6ft distancing and surface sanitation are estimated to reduce indoor disease transmission probability to a third of the transmission probability resulting from standard building ventilation practice.

## Introduction

A link is demonstrated between SARS-CoV-2 transmission efficiency and the indoor environment by correlating outdoor air temperature with an Infection parameter that characterizes the spread of Covid-19. Northern states in the US experienced surging virus transmission rates during colder weather while southern states were spared rampant disease spread. As heat and humidity forced southern US state citizens indoors, northern state citizens enjoyed comfortable outdoor conditions and decreased virus infection cases. The recent southern state infection surge has now moved north as northern state temperatures have increased above 70F, resulting in northern state citizens moving into sealed, inadequately ventilated, air conditioned building environments.

Today’s ventilation standards are based on odor, not health (1). And even in terms of odor-based ventilation design standards, buildings and homes throughout the US do poorly. Current building ventilation standards use an arbitrary criterion in which “only” 20 percent of the populace is *dissatisfied* with indoor air quality. We do not even reach that poor level of dissatisfaction as 80% of buildings fail to have only 20% air quality dissatisfaction (2).

You cannot smell healthy air. You cannot smell a virus. You cannot smell cold viruses, influenza viruses, nor the SARS-CoV-2 virus that is wreaking havoc throughout the world. The current debate among scientists of airborne versus direct and indirect contact infection modes is important, but equally important is determining how to stem the out-of-control growth of Covid-19. Buildings and homes must be changed from disease incubators to disease inhibitors by increasing fresh, filtered, and sanitized air flow into our buildings.

Today’s inadequate ventilation standards are estimated to ensure a 95% probability of a room’s occupants becoming infected with SARS-CoV-2 over the course of an 8 hour work day when occupied by an infectious person and three susceptible occupants. And those infected at school, work, and public gatherings will bring the virus home, where inadequate home ventilation and filtration ensures the likelihood of disease transmission among family members.

Reduction of disease transmission within buildings and homes requires increased fresh air flow rates (at least doubling to 40cfm per person) and improved air filtration (to at least MERV11 filtration). Carbon dioxide concentration monitoring of every indoor building space is the key to reducing indoor virus transmission rates. Carbon dioxide concentration is a direct measure of human respiration rates, and therefore, virus concentration in the indoor environment. Maintaining indoor carbon dioxide concentrations below 800ppm, equivalent to doubling today’s inadequate, odor-based ventilation rates, will reduce disease transmission rates below the limit required for decay of Covid-19 transmission.

## Current Covid-19 Disease Transmission Trends

Covid-19 was decaying in the US from May through mid-June 2020 as northern states gained control of infection spread through increased social distancing and decreased transmission efficiency. Figure 1 shows new case trends in the US over the past 4 months. As of mid-June, an alarming acceleration of new Covid-19 cases appeared in southern states.

**Figure 1.**
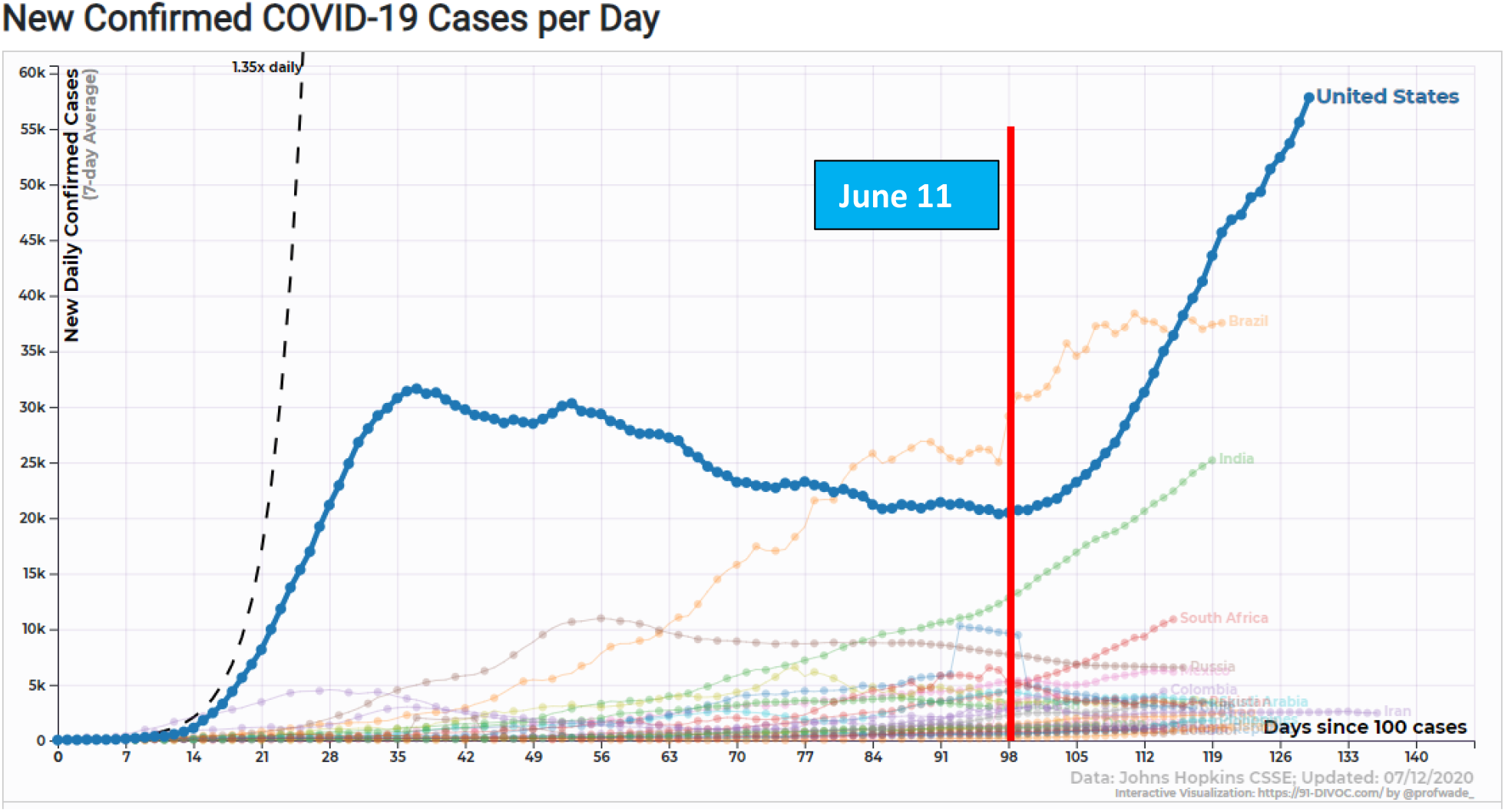
US new cases per day trend (7 day moving average). Data source: Johns Hopkins via 91-divoc.com; Professor Wade Fagen-Ulmschneider, University of Illinois.

A two-parameter Covid-19 prediction model has been developed based on a social distance index (SDI) and a disease transmission efficiency parameter (G). The appendices include formulation and assumptions used to formulate the model. The SDI is obtained from the University of Maryland/Maryland Transportation Institute and is based on anonymous cell phone and vehicle gps data. Covid-19 transmission efficiency describes the spread of the SARS-CoV-2 virus relative to transmission at a given SDI under “normal” human interactions (ie, no face masks, no 6ft distancing, hugging and handshaking allowed, etc) prior to virus containment efforts.

In mid-June, the disease transmission efficiency for the US increased abruptly while the social distance index remained relatively constant. The disease transmission efficiency parameter almost doubled from 0.1 to nearly 0.2 within a few days. Figure 2 is a plot of the infection parameter (IP, see Appendix B) as a function of SDI and G. Note that an IP value of ∼2.7 (the mathematical constant “e”) is a boundary in the Covid-19 prediction model separating infection growth and infection decay regions. An IP value of “e” produces constant infections per day and linear growth of total infection cases. The increase in IP from a value of approximately 2.7 to 4, due to the increase of virus transmission efficiency, causes a rapid escalation of new infection cases per day.

**Figure 2.**
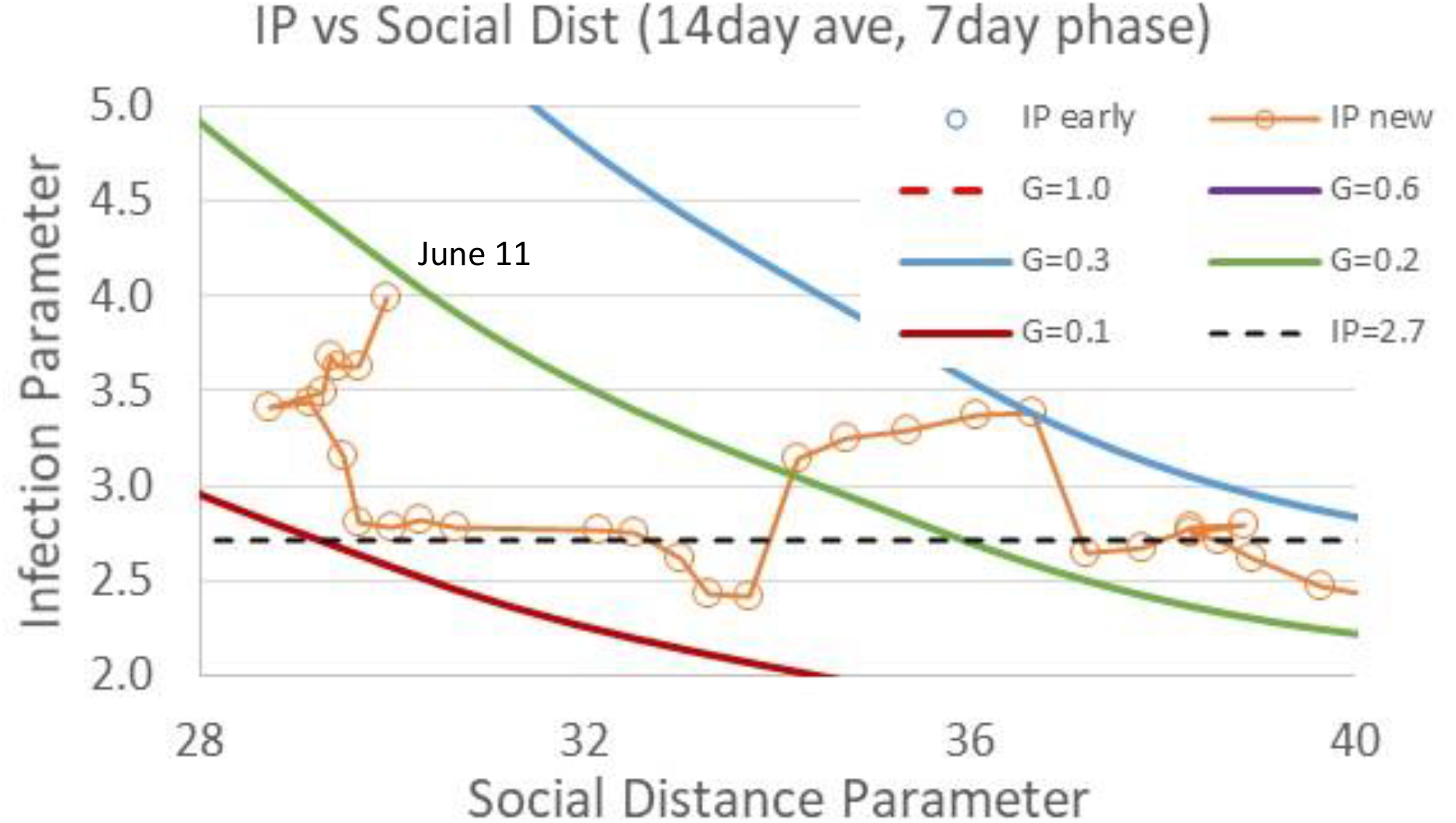
Infection parameter trends as a function of the Social Distance Index, SDI and the transmission efficiency, G. The last point with IP=4 at SDI=29.5 and G∼0.2 is for June 11 (SDI) and June 18 (IP) with a 1 week phase shift.

Figure 3 is a plot made in June comparing new daily case prediction trends versus actual data for the US. “Serious Isolation” assumes perfect isolation (G=0) of all uninfected and infected persons as of July 1 that results in decay of new Covid-19 cases over a 3 week period. A second path (“Fall Wave”) projected new Covid-19 cases assuming a social distance index of 30 with a 0.10 disease transmission efficiency parameter. The Fall Wave path is below the IP level of 2.7, resulting in a decay of new daily infections.

**Figure 3.**
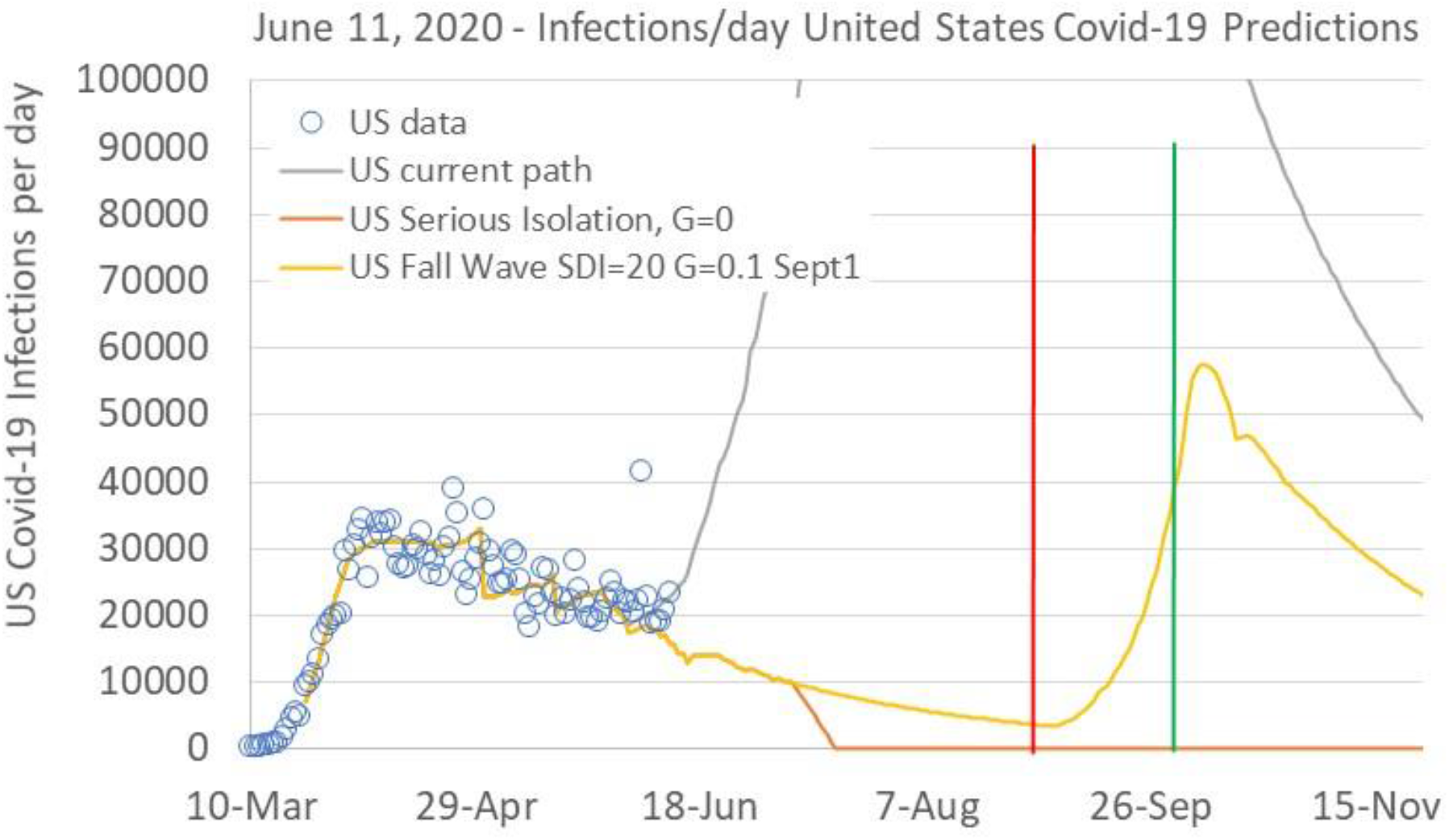
Mid-June US Covid infections per day predictions for the US for three infection paths.

The Fall Wave prediction in Figure 3 also assumes a decrease of SDI from 30 to 20 during the month of September while maintaining a transmission efficiency of 0.1. The decrease of SDI represents a scenario in which social interactions increase due to school re-openings and increased business and social activities. With the reduction of SDI to 20, IP increases above 2.7, resulting in an acceleration of new infection cases per day until SDI is increased or transmission efficiency is further decreased.

The third path plotted in the Figure 3 June prediction plot assumed an increase of transmission efficiency parameter to 0.22 while maintaining an SDI of 30. At that time, it was not clear if the increase of transmission efficiency was a transient occurrence that would fade or the beginning of a new trend. Figure 4 shows Figure 3 replotted 3 weeks later with actual data. The abrupt increase of the disease transmission efficiency parameter was not a brief excursion but instead a sustained transition in disease transmission characteristics.

**Figure 4.**
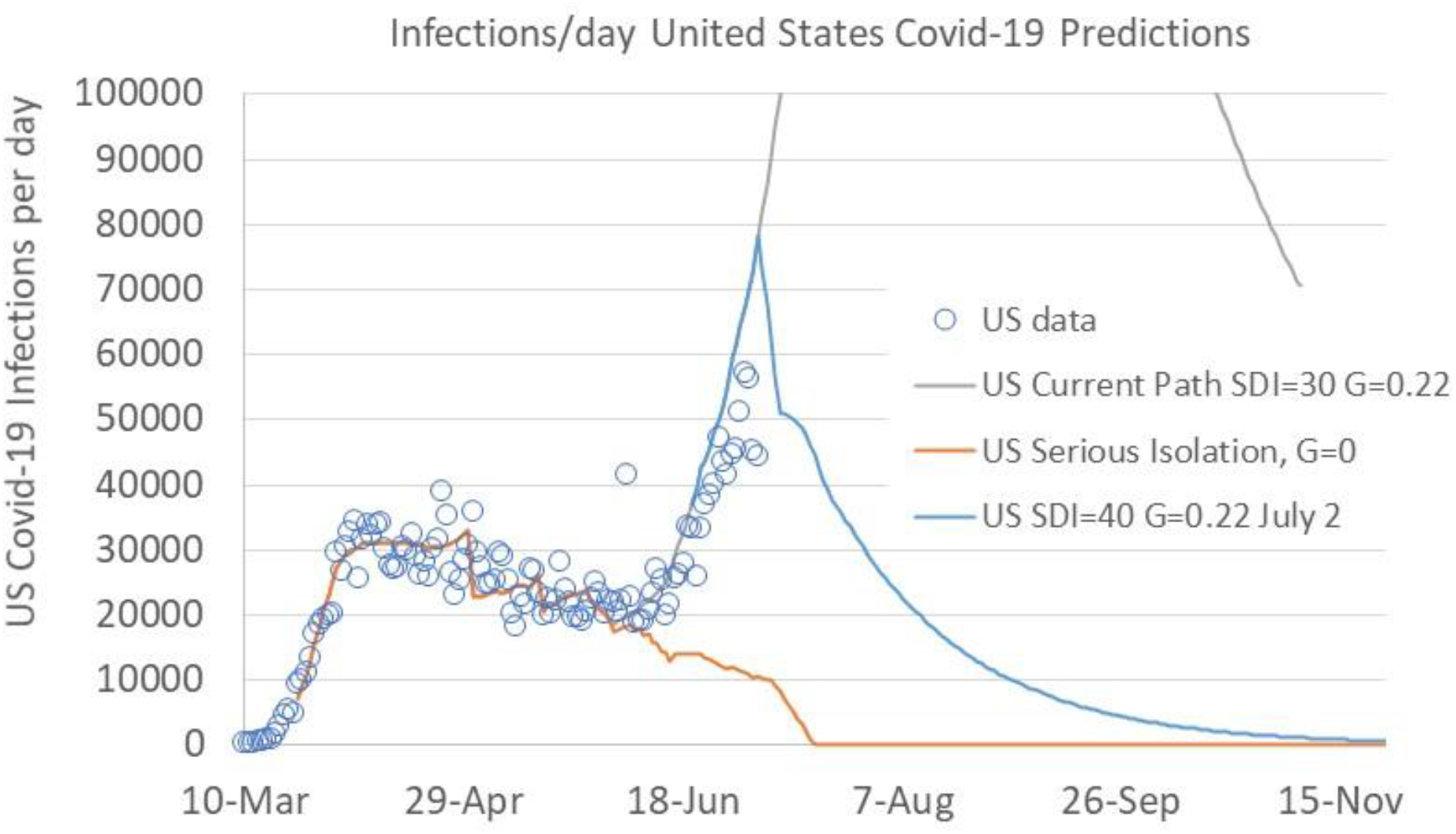
Early July US Covid-19 comparison between three prediction paths and actual Covid-19 daily new cases.

Infection growth is very sensitive to the infection parameter, and small increases above an IP value of 2.7 cause accelerated growth of new infection cases. Figure 4 shows a change of the social distance index to 40 with a disease transmission efficiency of 0.22 in early July would bring the infection parameter below 2.7, resulting in decreasing new infections. Figure 5 is an expanded version of Figure 4 with a series of plots showing increased social distancing occurring at different weeks in July. If unmitigated until August, an SDI of 30 with transmission efficiency of 0.22 would have grown to 400,000 new infections per day.

**Figure 5.**
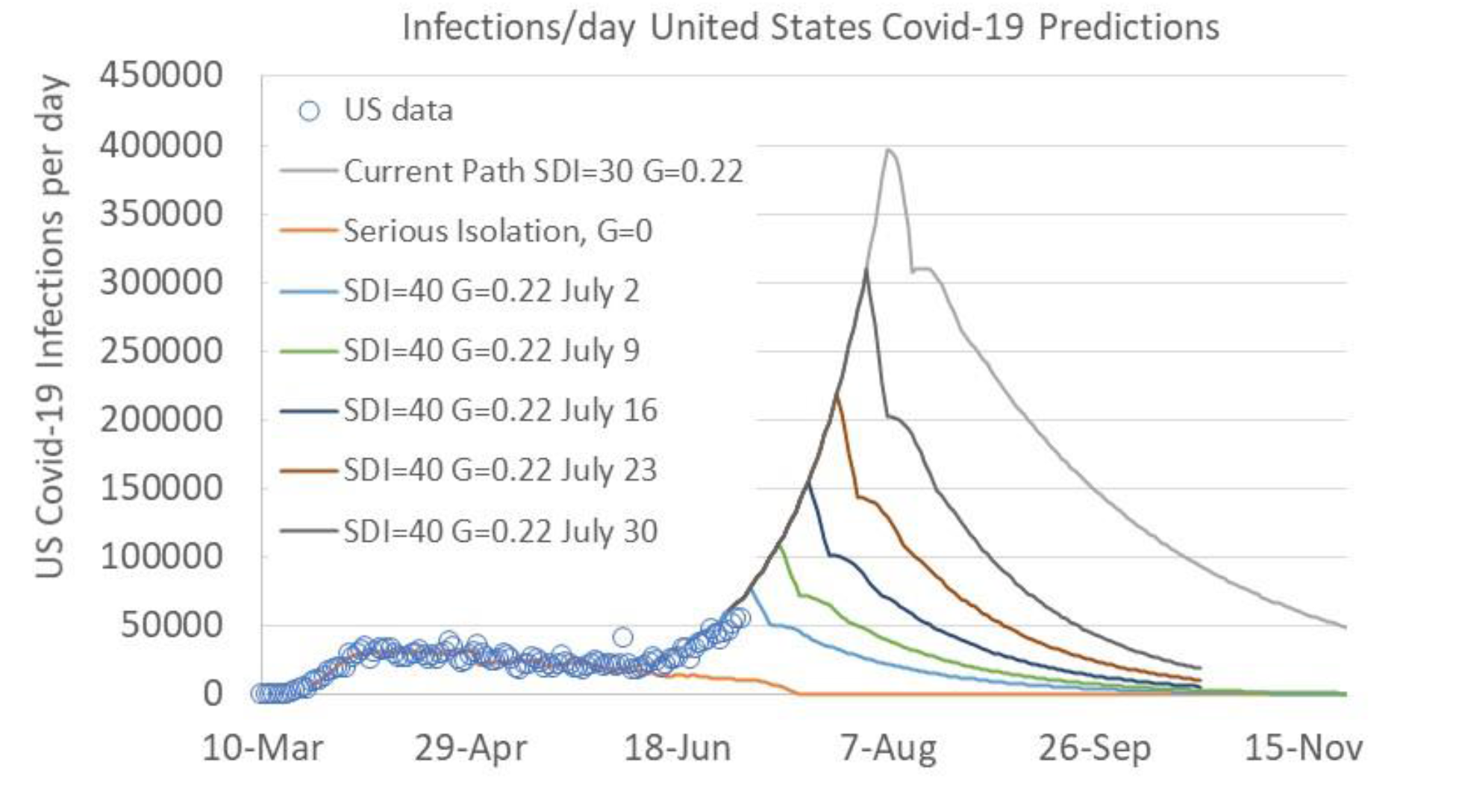
Early July Covid-19 daily new infections predictions showing impact of increased social distancing (SDI=40) enacted at different times during July.

Figure 6 is a plot of the Infection Parameter as of July 7, 2020, using the most recent posting of UMD SDI data. Since mid-June with extensive news coverage of uncontrolled, surging Covid-19 infections, the US populace has begun to increase social distancing. The disease transmission efficiency parameter is staying relatively steady at 0.19. A US social distance index greater than 36 is required to reach the infection parameter boundary of 2.7 if the disease transmission efficiency remains at 0.19.

**Figure 6.**
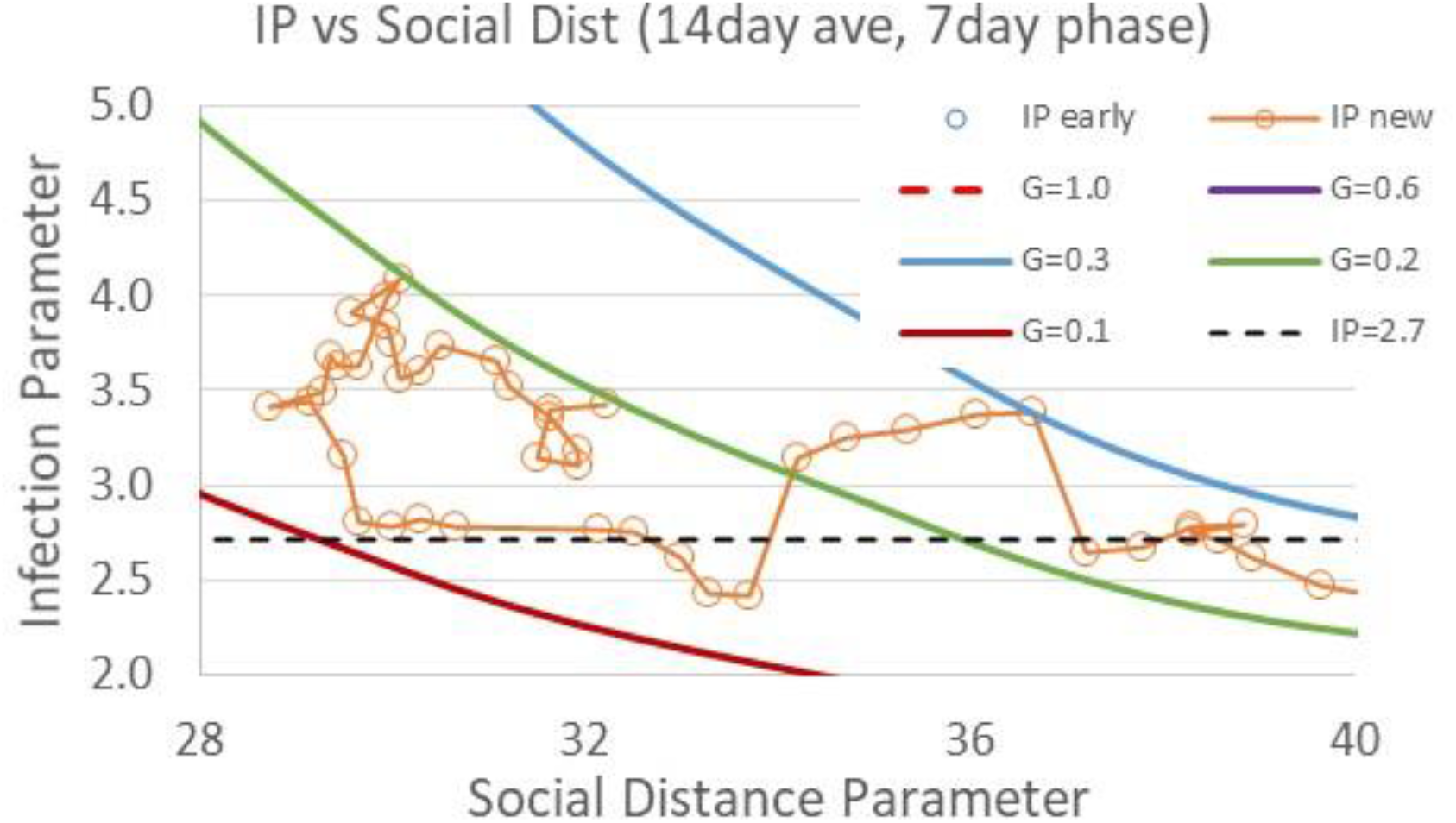
Infection Parameter (IP) trends including most current UMD SDI data as of July 7, 2020.

Figure 7 shows the infection parameter plotted versus the UMD social distance index since the emergence of the virus in the US in February. The US reached an average SDI of nearly 60 while some locations such as New York exceeded an SDI of 70. An SDI of 40 represents a reasonably open situation. Reduction of disease transmission efficiency is another key for reducing the infection parameter. The blue circle data points shown in Figure 7 are data points from the first few weeks of pandemic emergence in the US. These initial data points have been used to define a reference disease transmission efficiency parameter value of 1, representing a time before face masks, 6ft distancing, surface sanitizing, and other mitigation measures were in place. Appendices C and D describe correlation of the social distance index (SDI) to the infection parameter (IP), and defining the transmission efficiency parameter (G) with initial social distancing efforts in the US.

**Figure 7.**
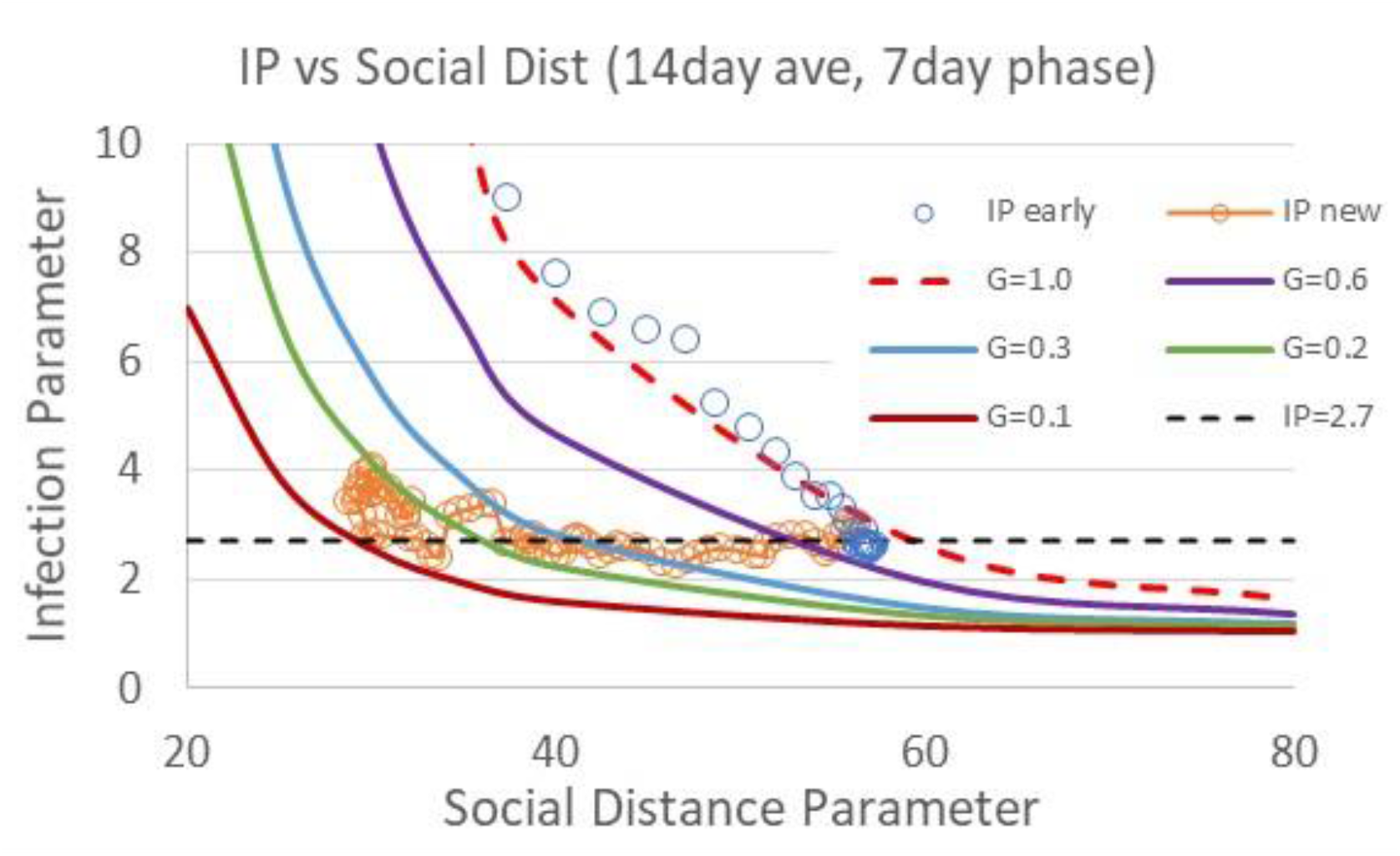
Expanded view of Infection Parameter (IP) trends showing early data (February-April) data as social distancing and business closures occurred, but prior to enactment of disease transmission efficiency efforts (eg, face masks, 6ft distancing, etc).

Figure 8 is an updated plot of new Covid-19 cases that incorporates the most current IP data trends from Figure 7. The “No Temp Effect” curve shows the trend in new Covid-19 cases assuming a constant SDI of 33 with a transmission efficiency parameter of 0.12 (that is, no abrupt increase). This case is a lower bound, assuming no change of today’s social interaction level while maintaining a low disease transmission efficiency that keeps the infection parameter well below the 2.7 infection growth limit.

**Figure 8.**
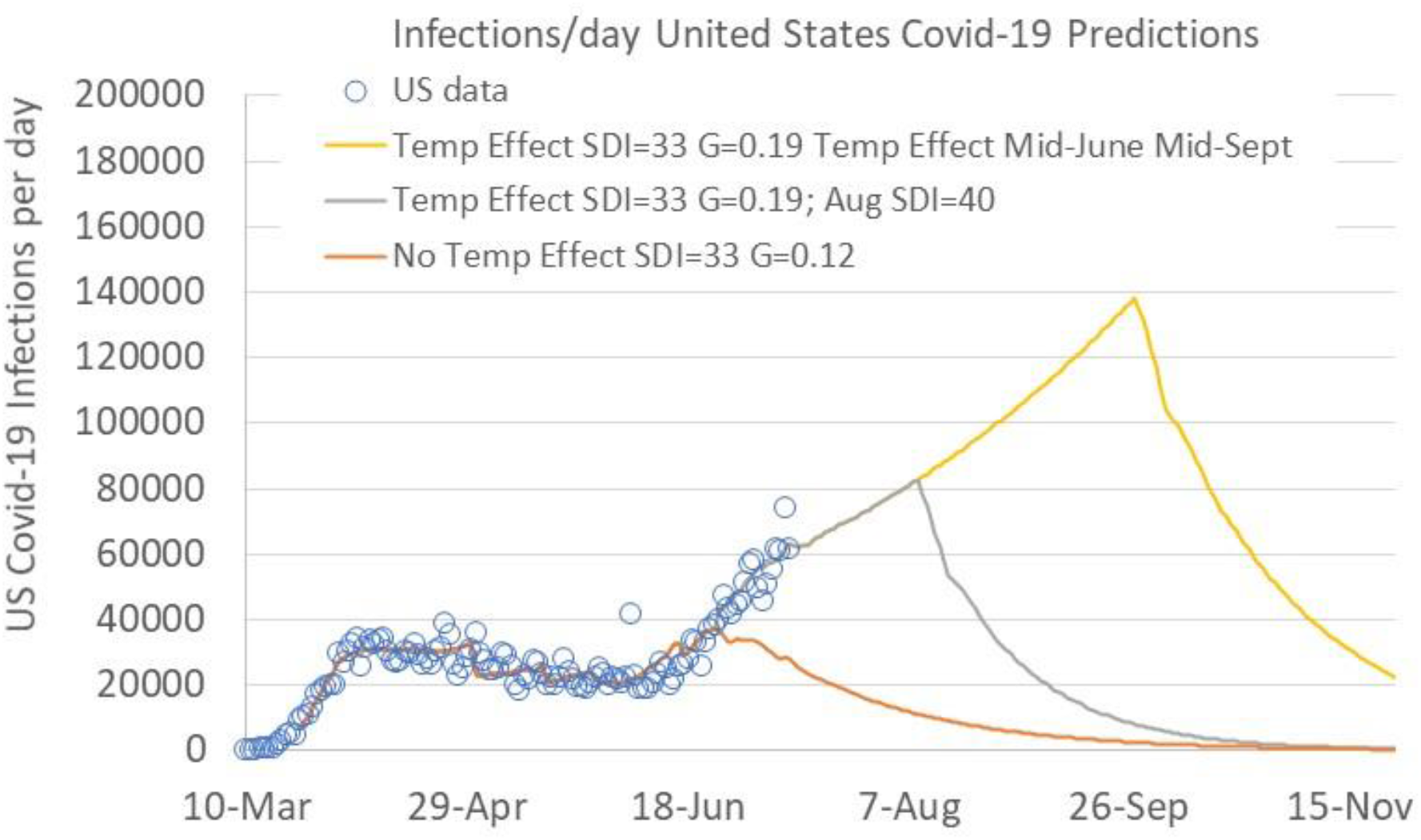
US Covid-19 predictions based on IP trends using current SDI and trasnmission efficiency (G) values. No temperature effect assumes G held constant at 0.12 with SDI=33, resulting in infection decay. Temperature effect paths assume increase of G to 0.19. The intermediate path assumes increased SDI to 40 in August, resulting in infection decay while the top path assumes no change of SDI with a drop in transmission efficiency in mid-September due to average US ambient temperature reduction.

Figure 8 shows two projected “Temp Effect” paths in which the social distance index is held at 33 with a disease transmission efficiency of 0.19. One case assumes US average outdoor temperatures above 70F keep the disease transmission parameter at 0.19 until mid-September while holding the SDI constant at The second case shows the effect of increasing social distancing to an SDI of 40 in early August. Based on the two Temp Effect cases in Figure 8, we should expect daily new infection cases to increase to 80,000 to 150,000 infections per day. Perhaps infections will be further dampened if people continue to increase the SDI as observed over the past few weeks.

The next section describes infection parameter trends in individual states, followed by a section that identifies the movement of people into buildings when outdoor temperatures are below 50F or above 70F as a primary cause for increased Covid-19 disease transmission efficiency. We recommend steps for decreasing the transmission efficiency factor based on increased fresh air ventilation, improved indoor air filtration, and integration of UVGI (ultraviolet germicidal irradiation) to building ventilation systems. We estimate these steps will decrease the probability of infection to a third of that for a building with industry standard ventilation (ASHRAE 62.1/62.2) levels, resulting in an overall decrease of the Covid-19 disease transmission efficiency parameter.

## State Infection Parameter Trends

Figures 9-14 show Infection Parameter (IP) plots for New York, Illinois, Ohio, Michigan, Minnesota and California, similar to Figure 7 for the US. New York (Figure 9) closed businesses and schools in March and April, with a social distance index of 70 reached in April. New York enacted and encouraged face masks, 6ft spacing, sanitation, as well as closing mass transit systems. These actions decrease the disease transmission efficiency parameter as seen in downward movement of the infection parameter at a given social distance index. As New York relaxed social distancing, they were able to keep the infection parameter well below the 2.7 IP boundary that divides infection growth and infection decay regions. New York continues to maintain a relatively high SDI in the 40s, however, similar to other locations, the disease transmission efficiency has increased over the past few weeks without any significant change to SDI or disease transmission reduction measures.

**Figure 9.**
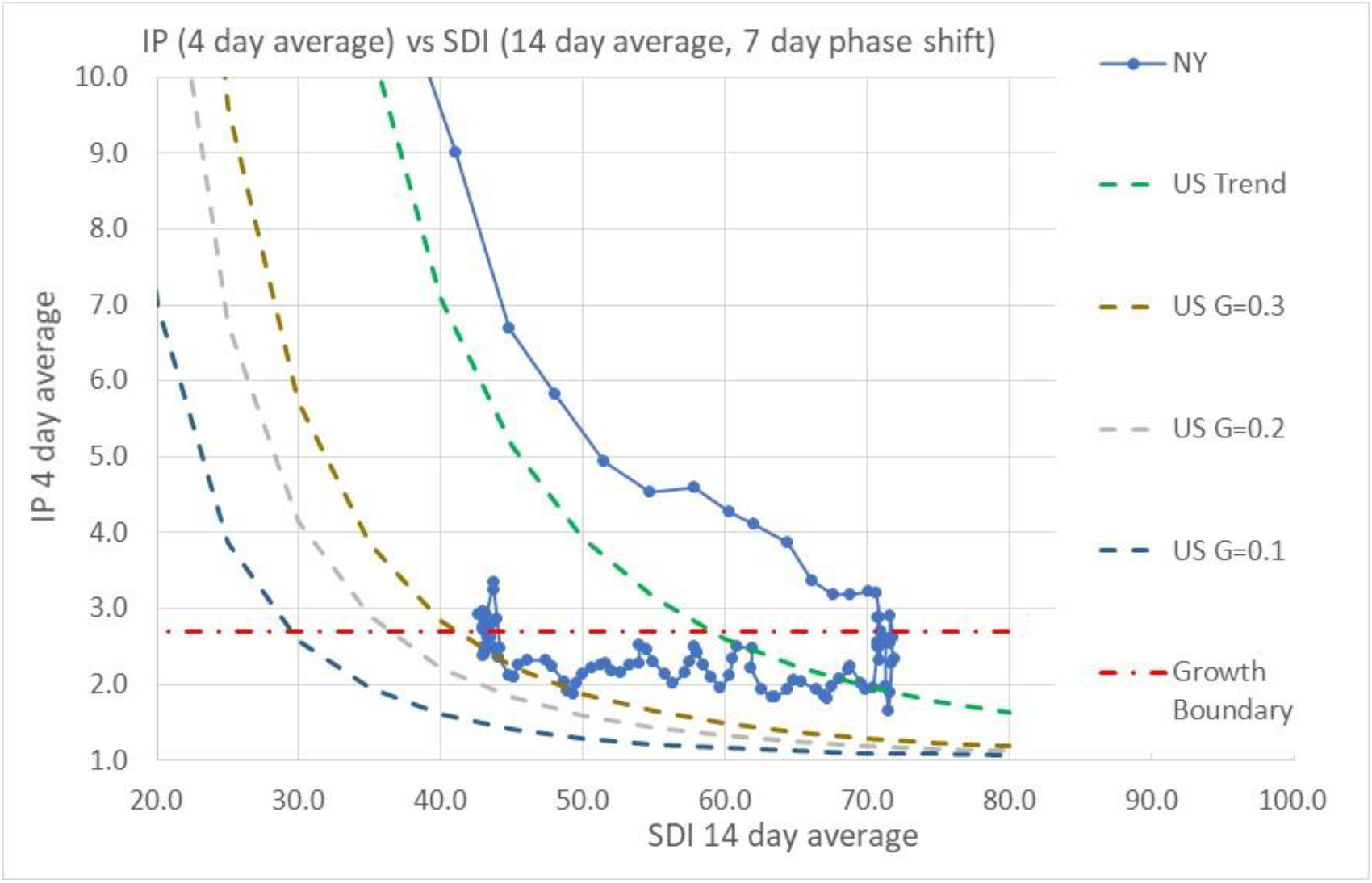
New York Infection Parameter (IP) trends as a function of the UMD Social Distance Index and Covid-19 disease transmission efficiency parameter, G.

**Figure 10.**
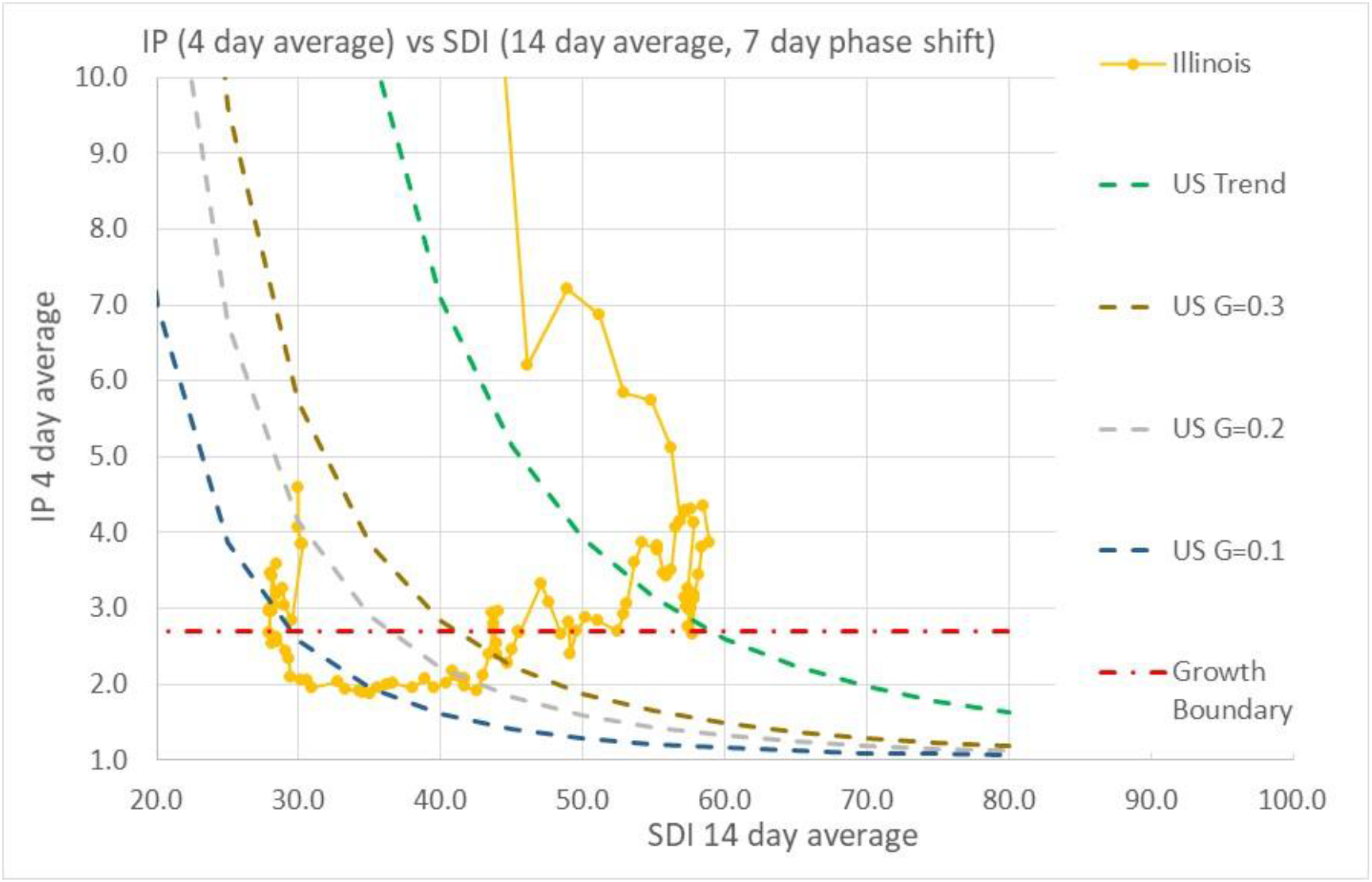
Illinois Infection Parameter (IP) trends as a function of the UMD Social Distance Index and Covid-19 disease transmission efficiency parameter, G.

**Figure 11.**
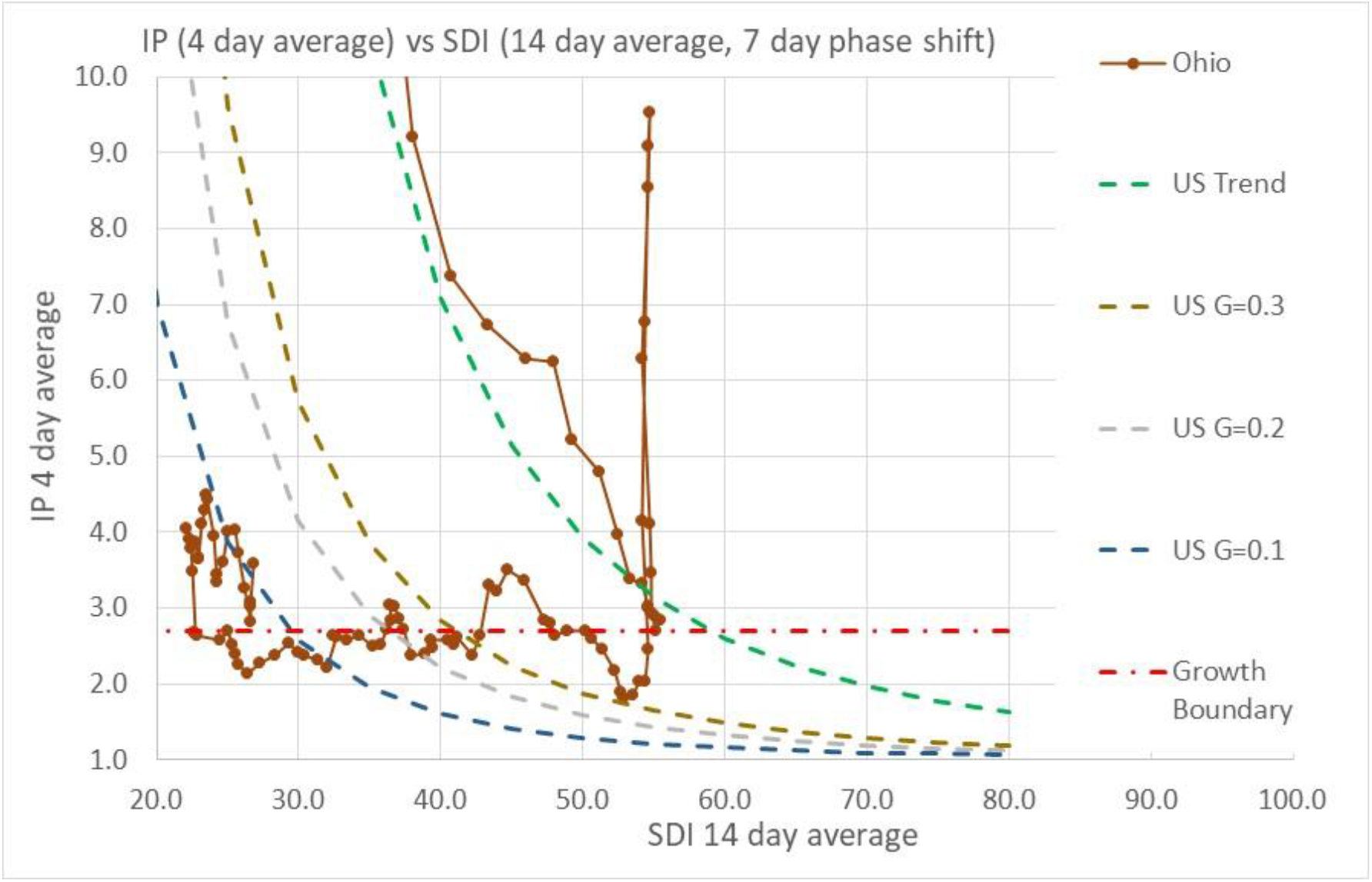
Ohio Infection Parameter (IP) trends as a function of the UMD Social Distance Index and Covid-19 disease transmission efficiency parameter, G. Note that the IP “spike” at SDI∼55 is due to an outbreak in a confined environment (prison).

**Figure 12.**
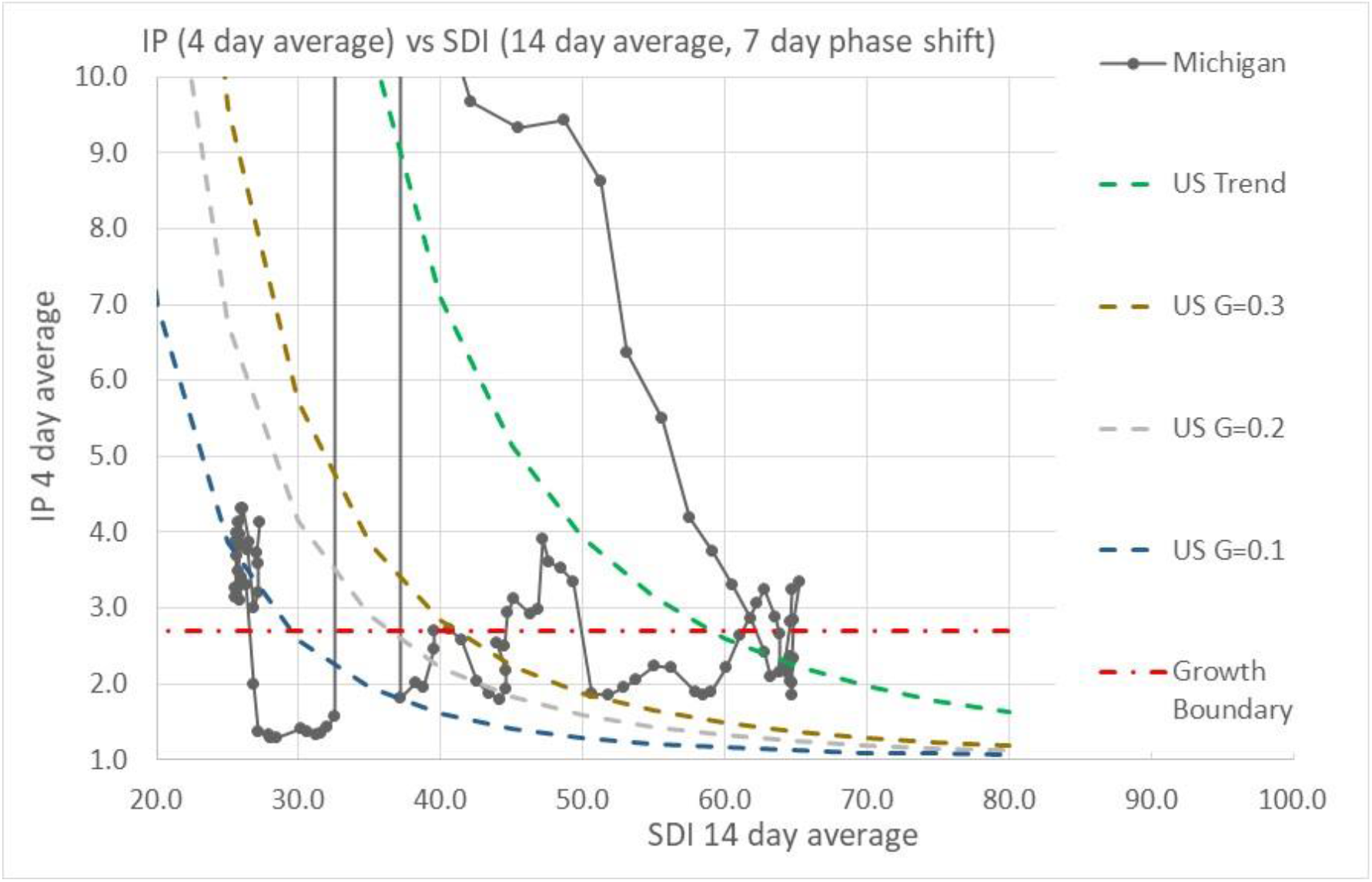
Michigan Infection Parameter (IP) trends as a function of the UMD Social Distance Index and Covid-19 disease transmission efficiency parameter, G. Note that the IP “spike” at SDI∼35 is due to a change in disease case reporting by the State.

**Figure 13.**
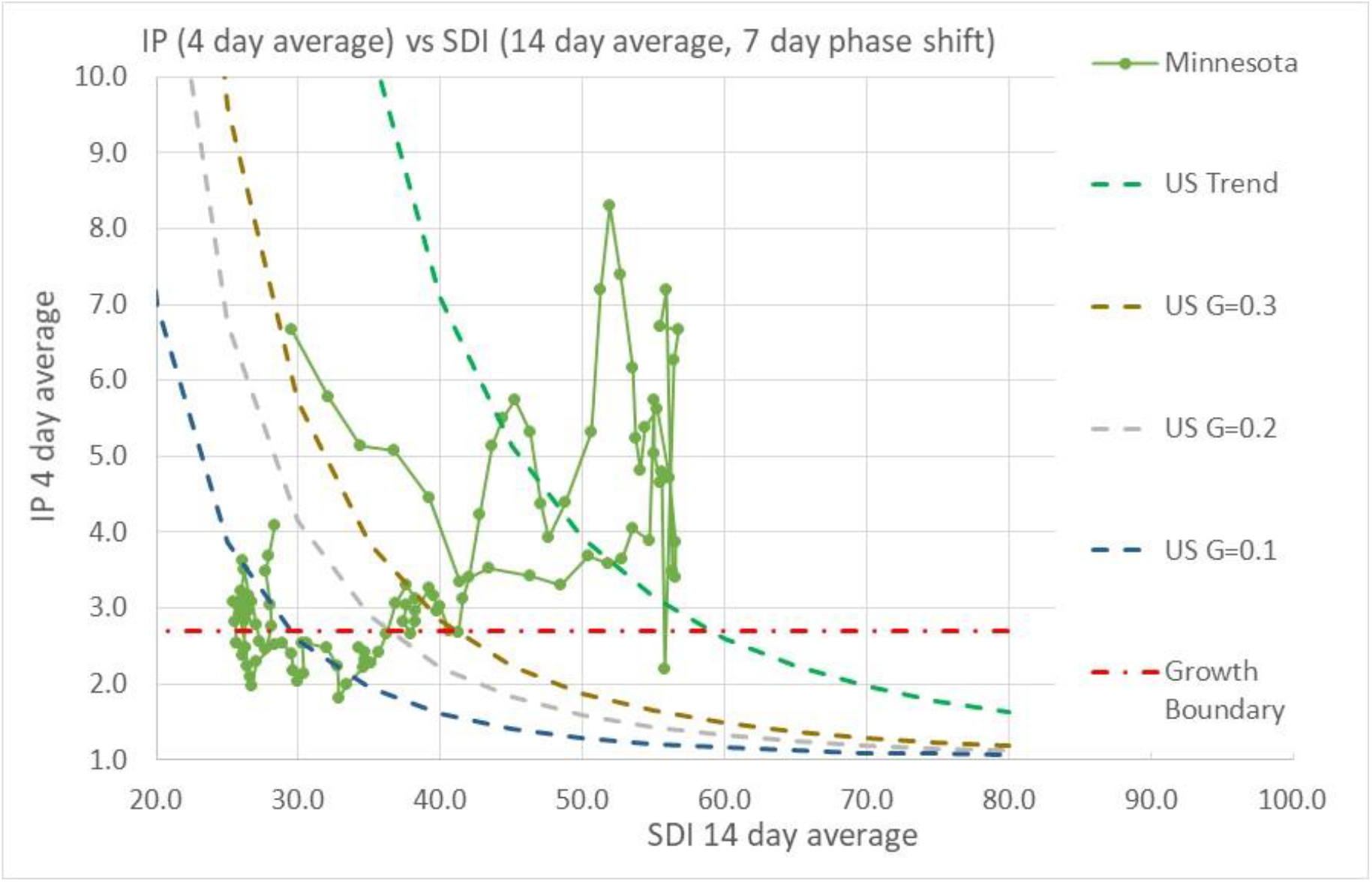
Minnesota Infection Parameter (IP) trends as a function of the UMD Social Distance Index and Covid-19 disease transmission efficiency parameter, G. Note that Minnesota case reporting was erratic during early stages of Covid-19.

**Figure 14.**
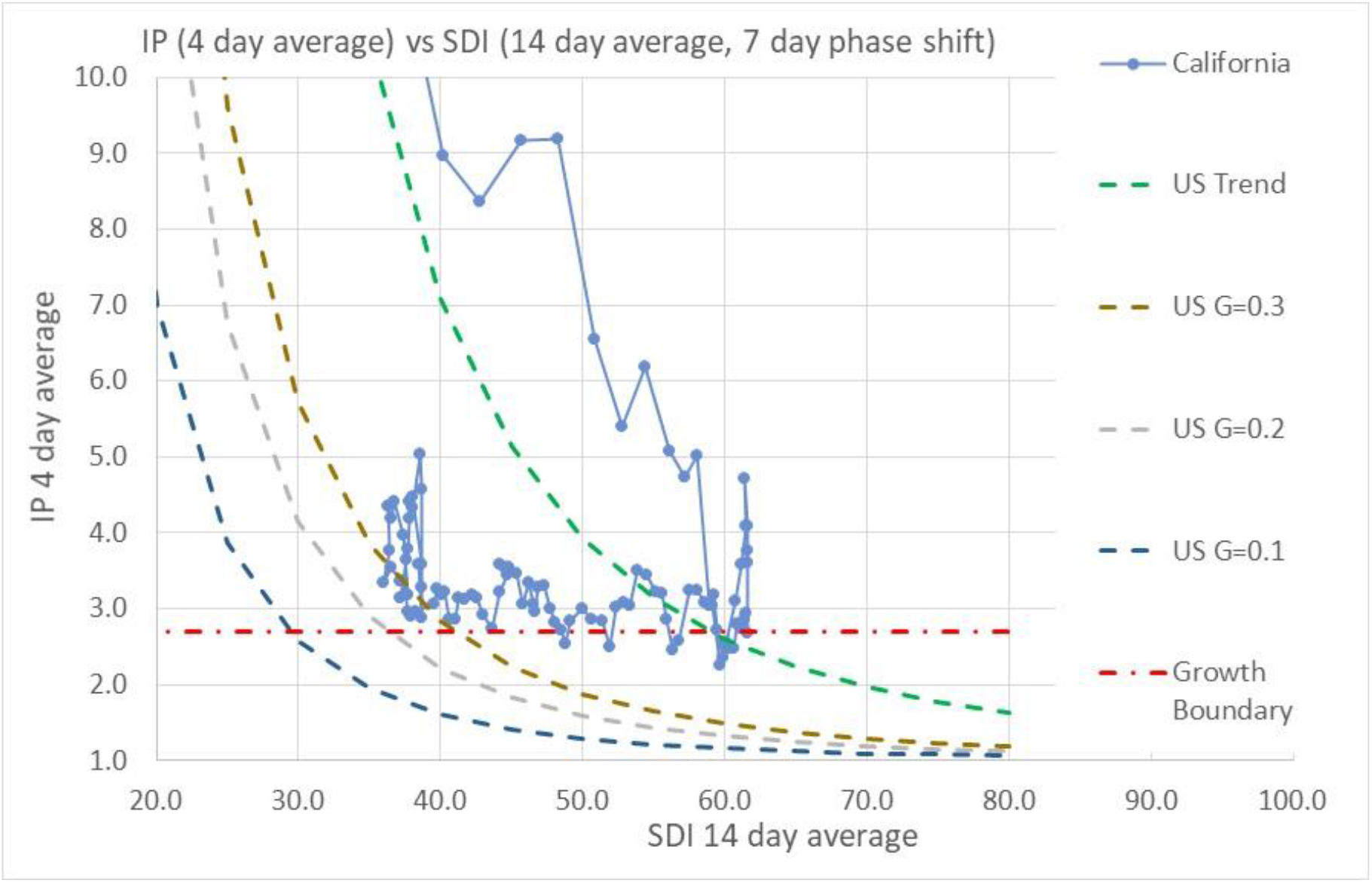
California Infection Parameter (IP) trends as a function of the UMD Social Distance Index and Covid-19 disease transmission efficiency parameter, G.

Illinois followed a similar path as New York as shown in Figure 10. Illinois had difficulty reaching as high of an SDI as New York, with a maximum SDI of 60. Even though social distancing measures were being relaxed in April and May, resulting in lowered SDI, Illinois continued pushing disease transmission efficiency measures, such as face masks, resulting in a drop of the infection parameter below 2.7. Over the past two weeks, a significant jump of transmission efficiency has increased the infection parameter to levels above the 2.7 boundary, with Illinois now experiencing accelerated daily infection growth.

Ohio (Figure 11), Michigan (Figure 12), and Minnesota (Figure 13) followed similar trends as Illinois, with a steadily decreasing infection parameter as social distancing was increased and decreasing disease transmission efficiency with face masks, 6 ft distancing and other transmission inhibiting actions. As of the past two weeks, all three states have experienced a similar jump in infection parameter without any significant change in social distancing. Note that Minnesota’s Covid-19 case reporting was erratic during the early days of case reporting, causing some noise in data during that time.

California (Figure 14) also shows a similar trend as the other states, although California has continually had difficulty controlling the virus. Unlike the other states discussed, California has not reached a point in which the infection parameter has consistently been held below the 2.7 limit.

Figures 15 and 16 display infection parameter trends for Illinois and Michigan (Figure 15) and Ohio and Minnesota (Figure 16) versus date. All infection parameter plots show a similar abrupt increase of infection parameter during the recent past as warmer weather moved into northern states. Figure 17 is a plot of the US infection parameter trends versus date, with a less abrupt increase of infection parameter that reflects geographical averaging of warm weather over a large, diverse area.

**Figure 15.**
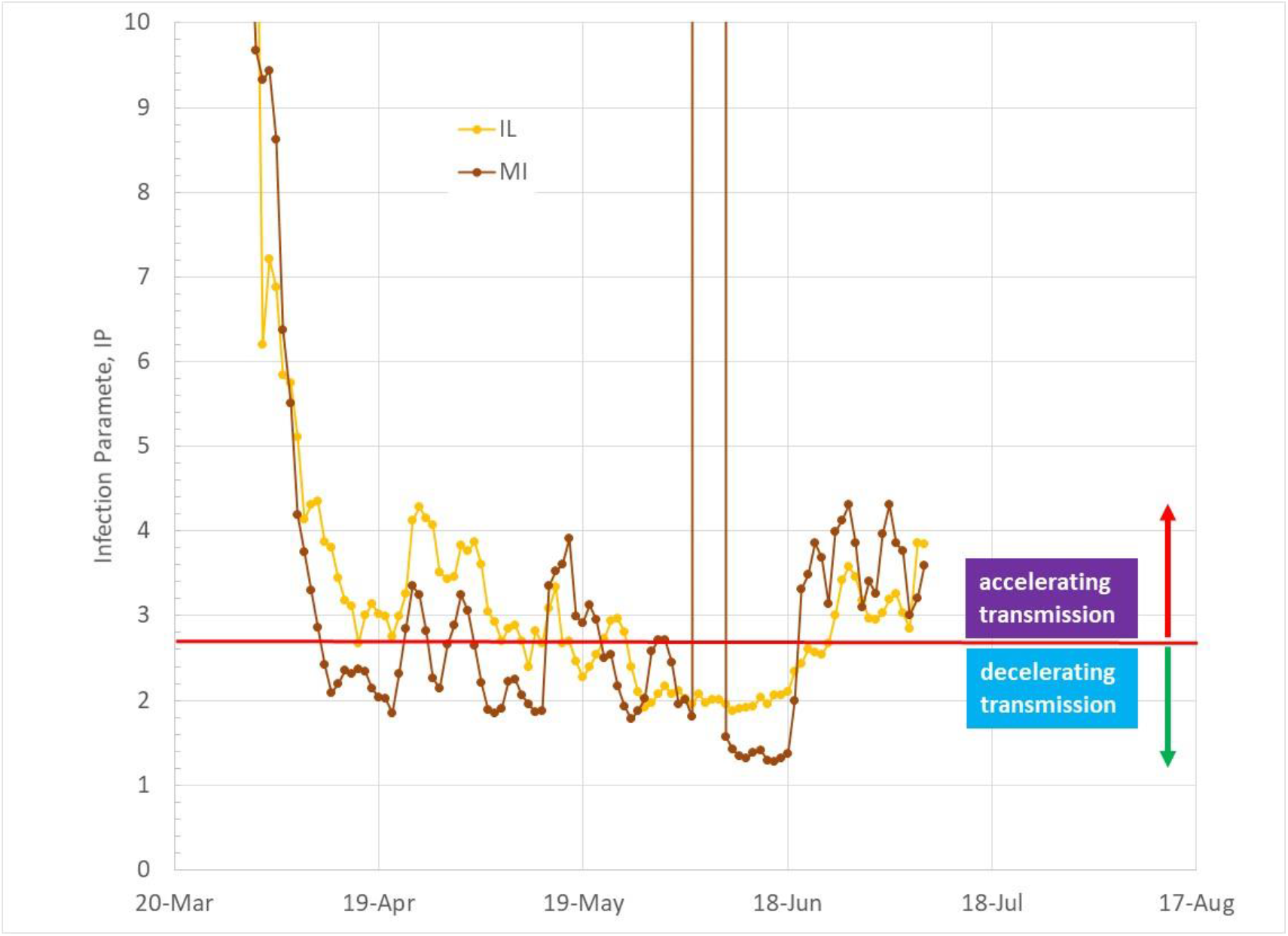
Illinois and Michigan Infection Parameter (IP) trends as a function of date.

**Figure 16.**
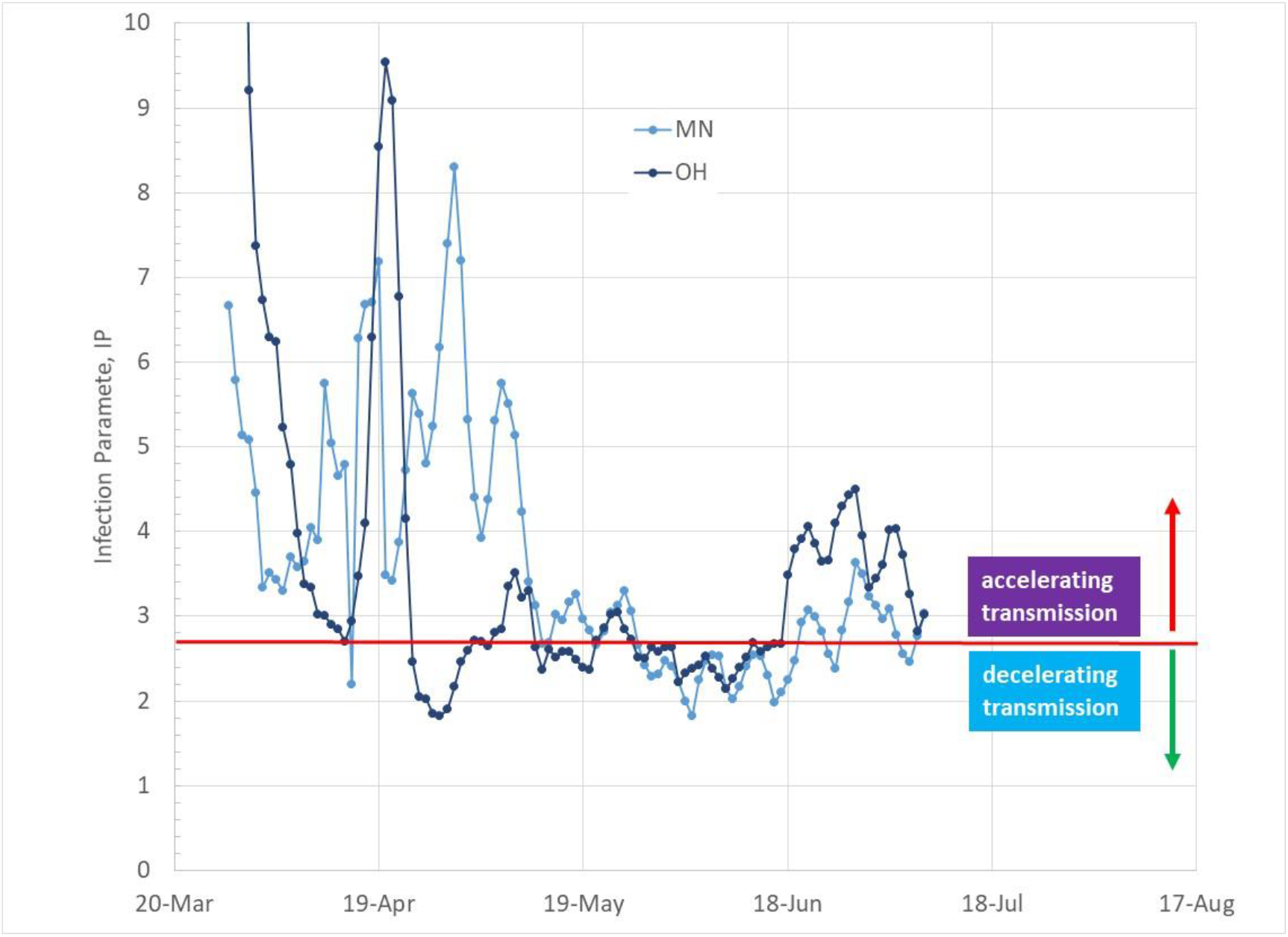
Ohio and Minnesota Infection Parameter trends as a function of date.

**Figure 17.**
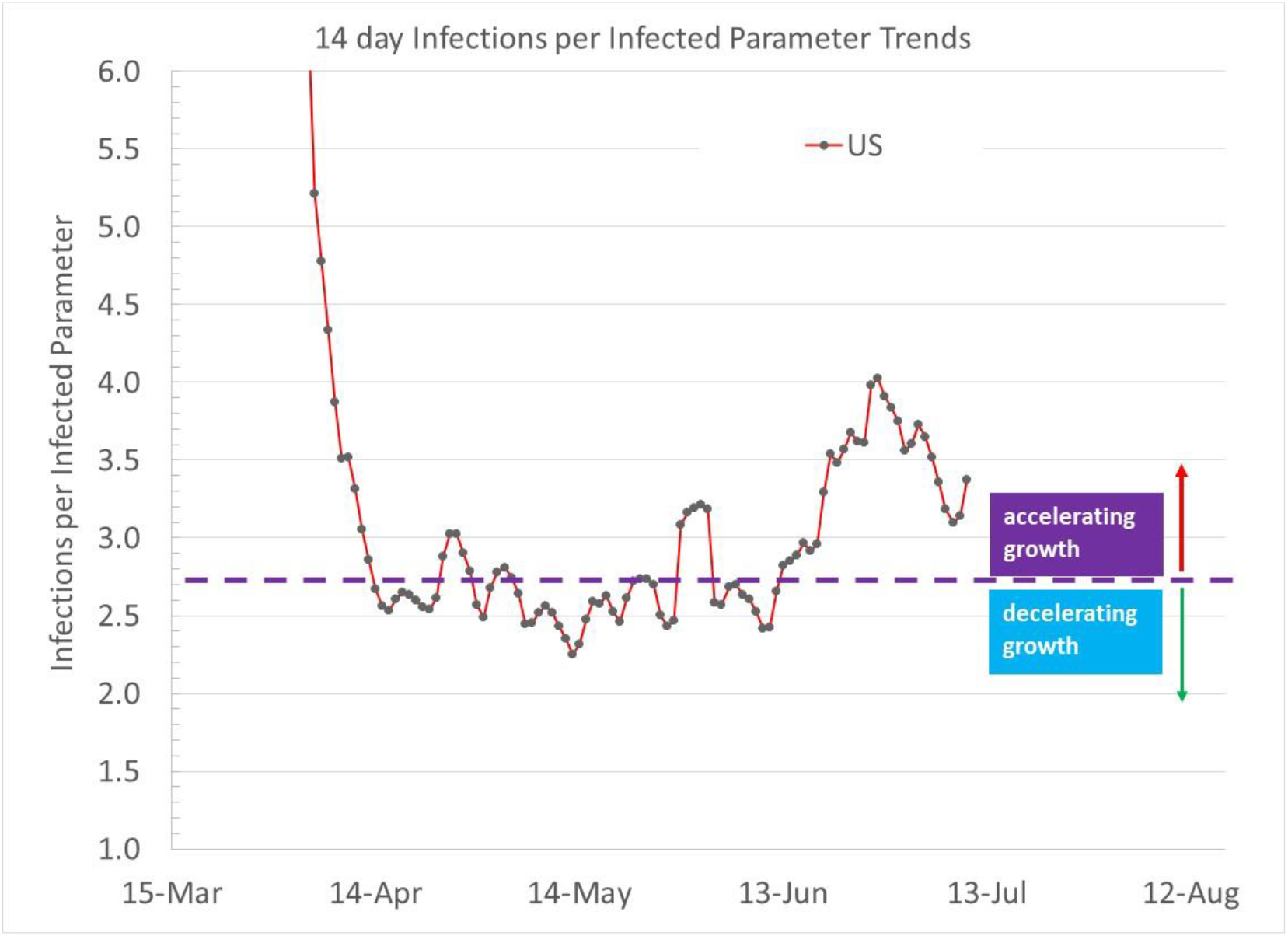
US Infection Parameter trends as a function of date.

## Outdoor Temperature and Comfort Conditioning

Northern states experienced much higher SARS-CoV-2 transmission rates than southern states during the early outbreaks in the US during February and March. As the US shutdown and enacted social distancing in March 2020, outdoor weather also changed from winter to spring in the north, with people increasing outdoor activities as well as opening their windows and doors. Southern state citizens, during this time, were enjoying outdoor activities and opening homes to fresh air during the late winter and early spring.

We were lulled into thinking Covid-19 had been defeated by May 2020 as heavily infected northern states reduced infection rates. SARS-CoV-2, unfortunately has not shown a sensitivity to climate (temperature and humidity) as do other respiratory viruses. Decreased virus survivability has been attributed to the summer decline of the 1918 influenza pandemic.

Another major difference between the 1918 influenza pandemic and today’s Covid-19 pandemic is comfort conditioning. No homes or buildings had air conditioning in 1918 (yes, a few buildings had AC; beginning with the 1902 installation of a cooling system in the Wilhelms-Sackett Lithography Co in Brooklyn, followed by the New York Stock Exchange). During summer conditions in the early 1900s, building ventilation was maximized by people opening windows and doors, working outside, teaching in outdoor classrooms, and sleeping outdoors (eg, “sleeping” porches).

Today, people move indoors when conditions are uncomfortable outside with minimal ventilation, causing contagion concentration increases indoors. Figure 18 shows daily comfort conditioning energy data from [13 identical homes](https://www.buildequinox.com/files/CERV/VermodCERVReport06262016.pdf) occupied by non-identical people (1 to 5 occupants per home). The data, collected over a 2 year period, shows comfort conditioning energy is a minimum during the “swing seasons” when outdoor temperatures range from 50F to 70F. Within the 50 to 70F outdoor temperature range, most people transition from heating to cooling in the spring, and vice versa in the fall.

**Figure 18.**
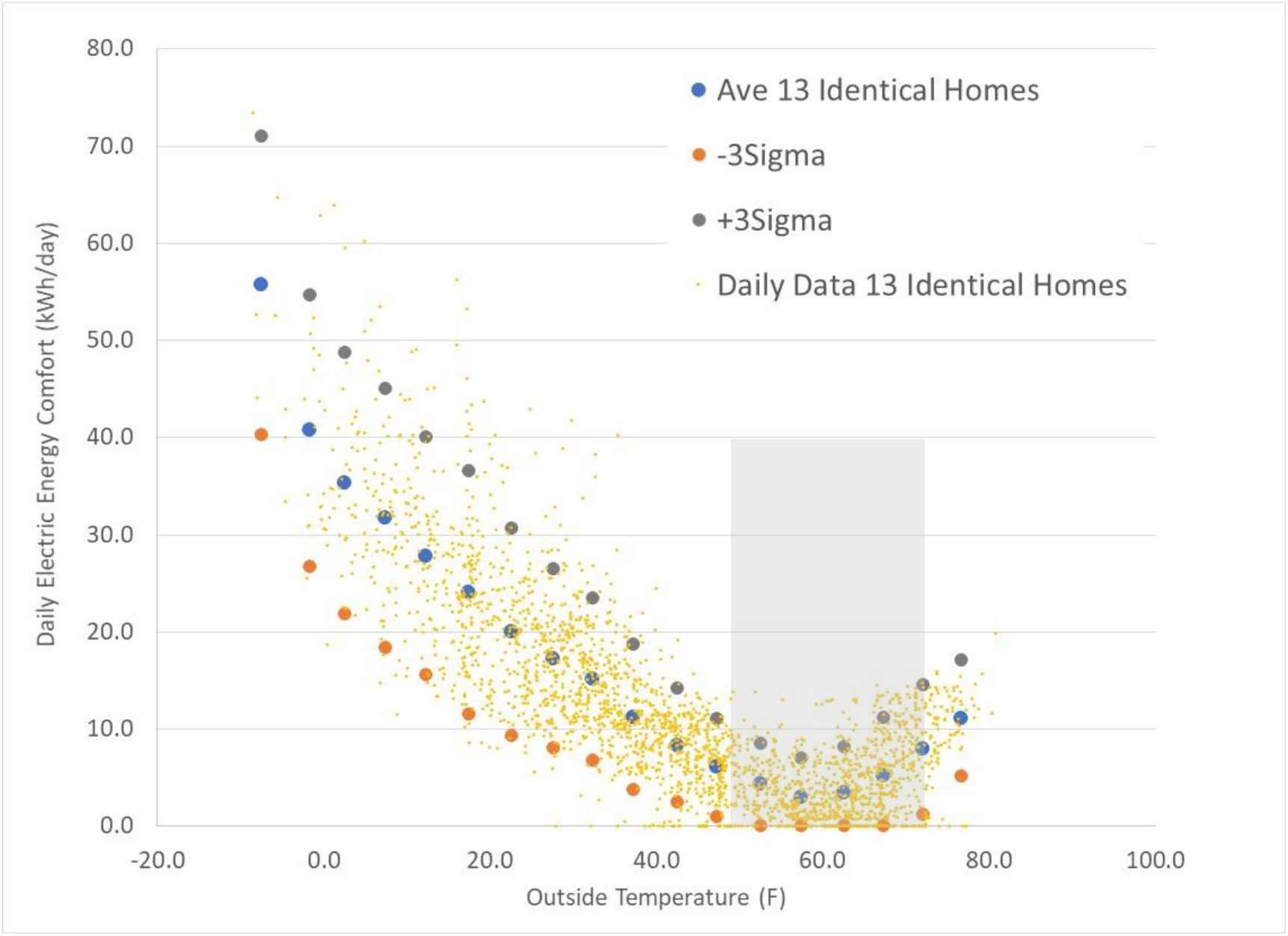
Daily comfort conditioning energy data for 13 identical homes over a 2 year period. Comfort conditioning energy is a minimum between 50F and 70F when many homes are opened to nice outdoor conditions.

Figures 19-24 show state Infection Parameter (IP) trends plotted against outdoor ambient temperatures. Daily infection parameter values and daily average outdoor temperatures are 14 day moving averages that characterize the 14 day infectious period of Covid-19. The average temperature data is shifted one week earlier from the average infection parameter to account for the disease incubation period, consistent with the averaging and shift between IP and SDI as discussed in Appendix C. That is, today’s infections are related to weather and social interactions that occurred a week ago.

**Figure 19.**
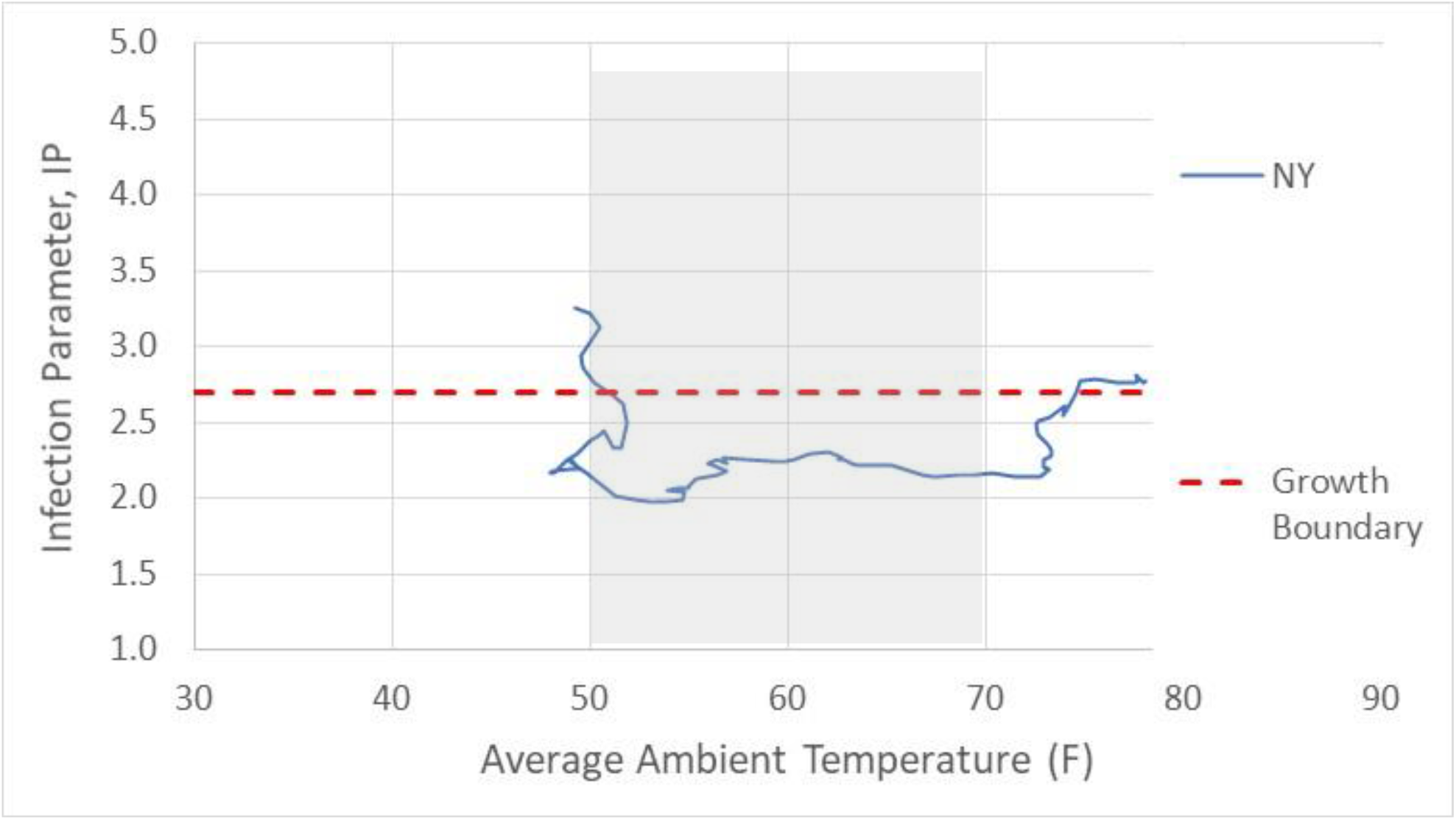
New York Infection Parameter trends with outdoor daily average ambient temperature (New York City).

**Figure 20.**
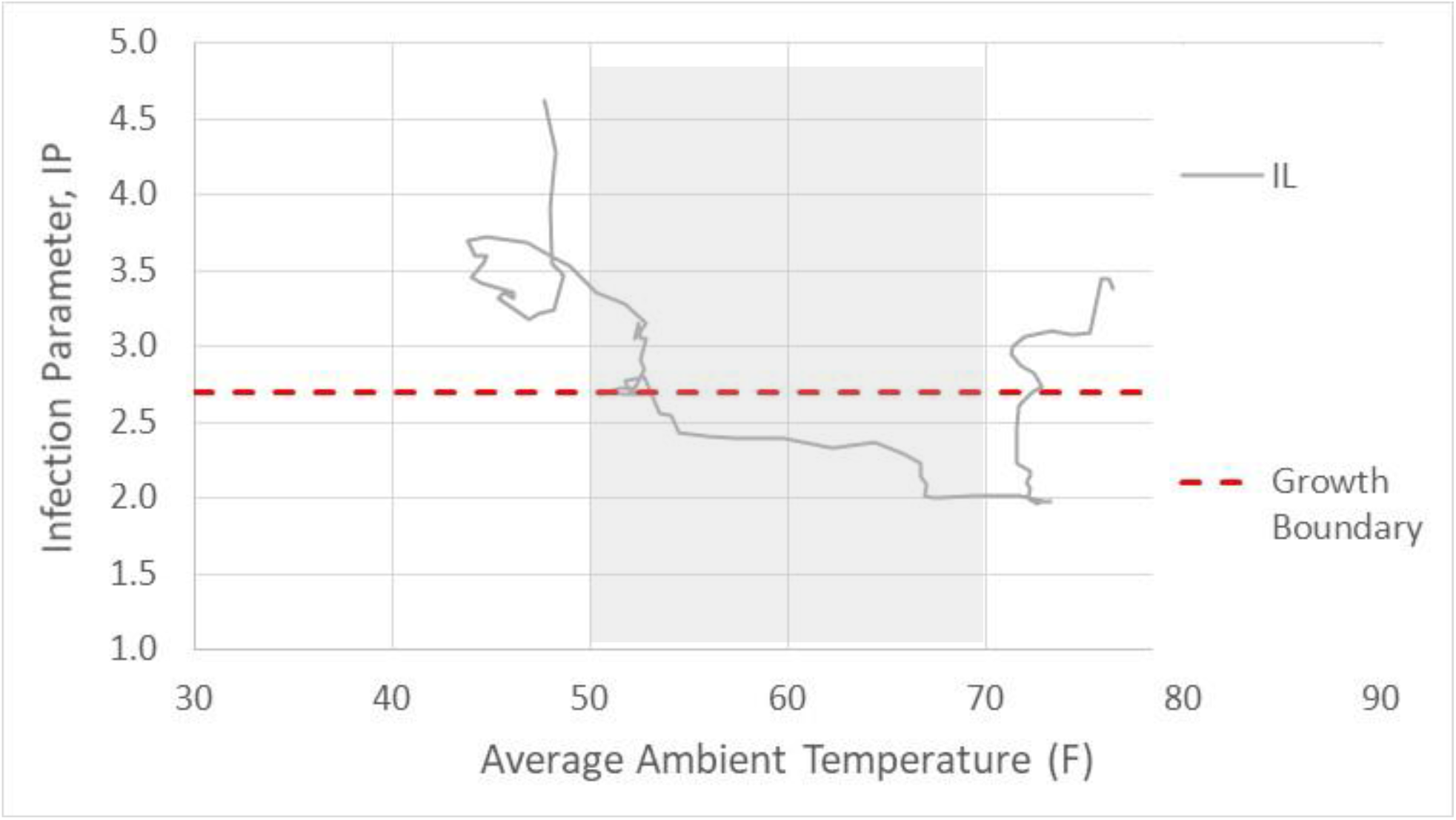
Illinois Infection Parameter trends with outdoor daily average ambient temperature (Chicago).

**Figure 21.**
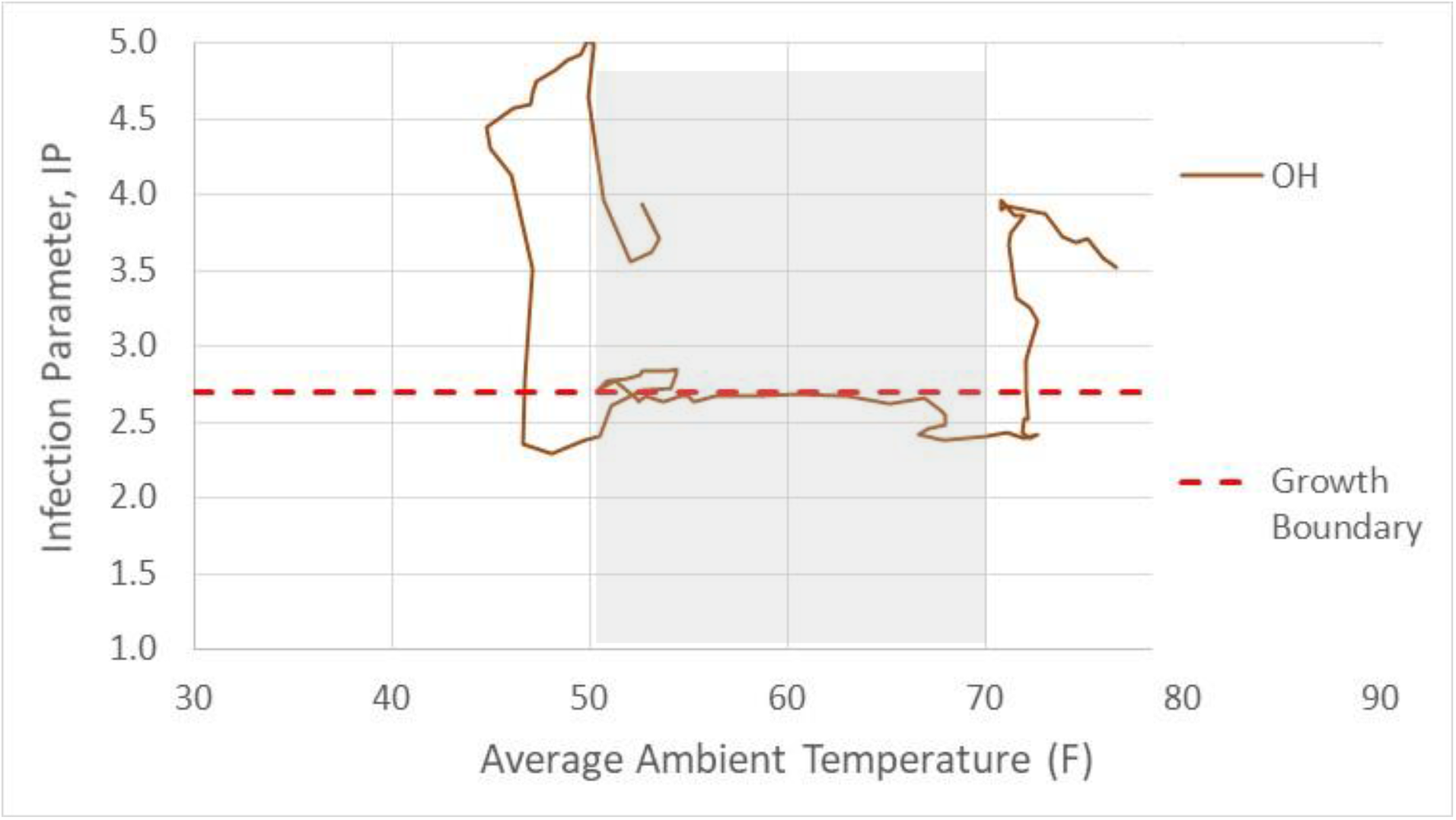
Ohio Infection Parameter trends with outdoor daily average ambient temperature (Columbus).

**Figure 22.**
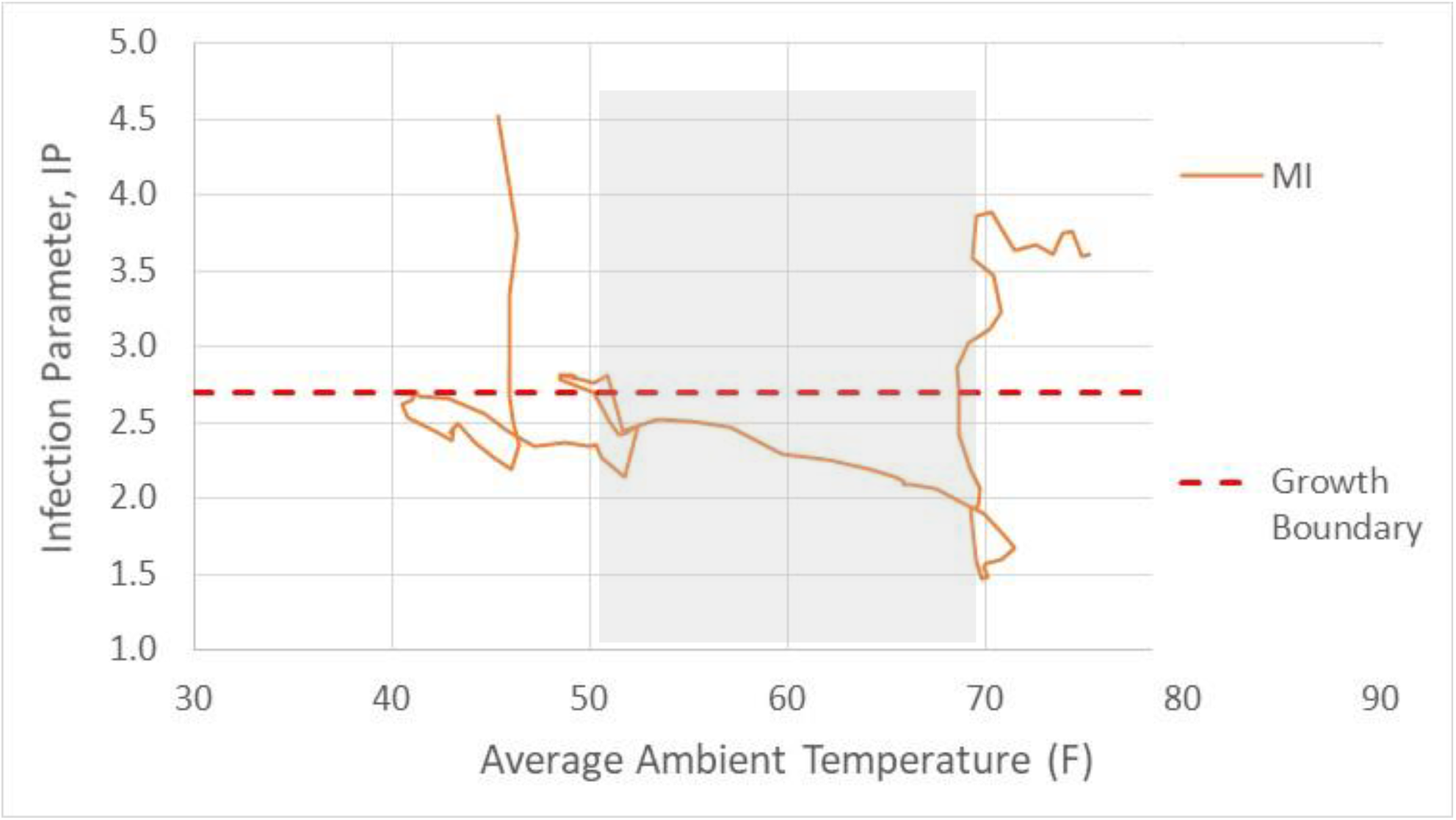
Michigan Infection Parameter trends with outdoor daily average ambient temperature (Detroit).

**Figure 23.**
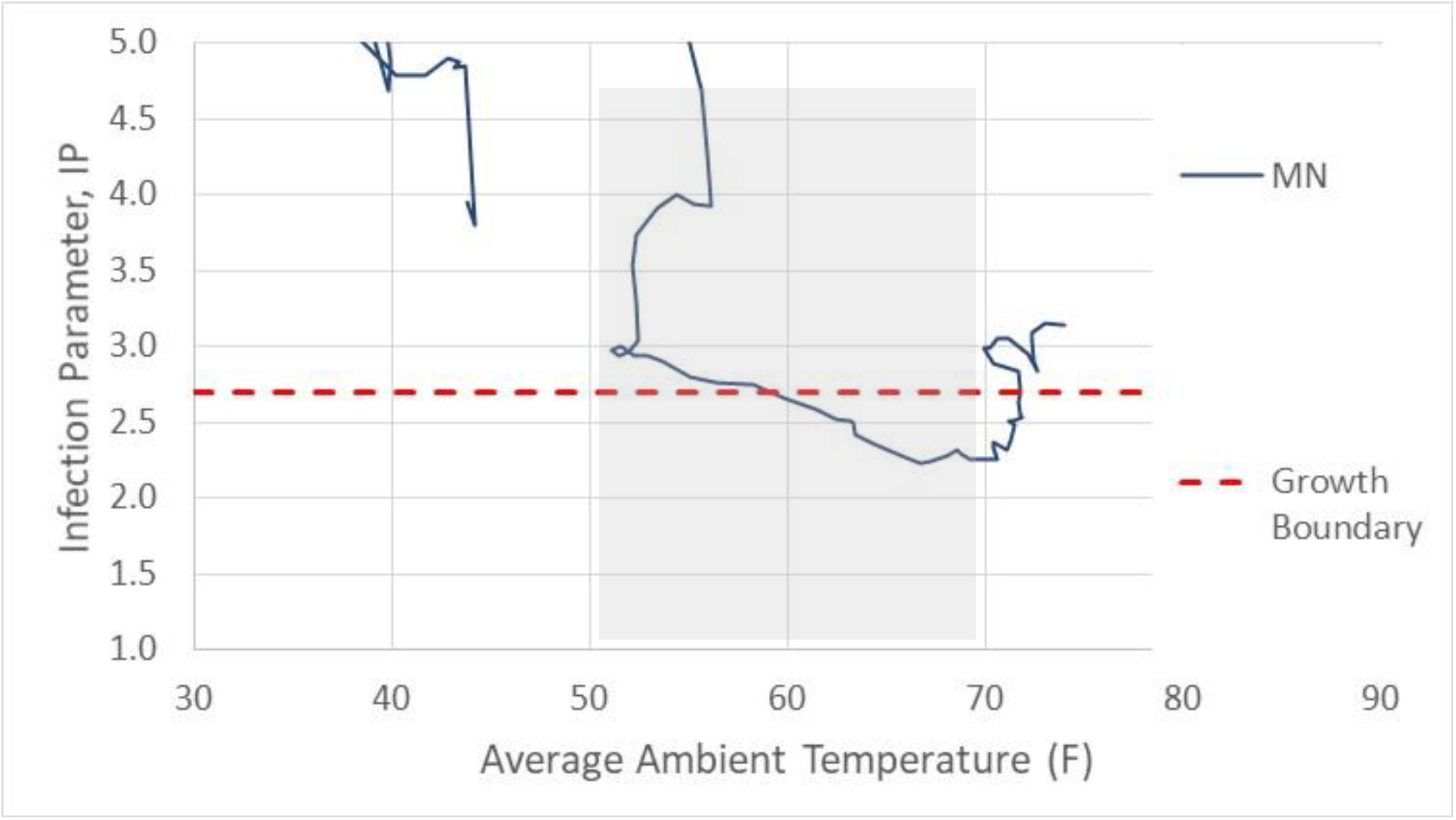
Minnesota Infection Parameter trends with outdoor daily average ambient temperature (Minneapolis).

**Figure 24.**
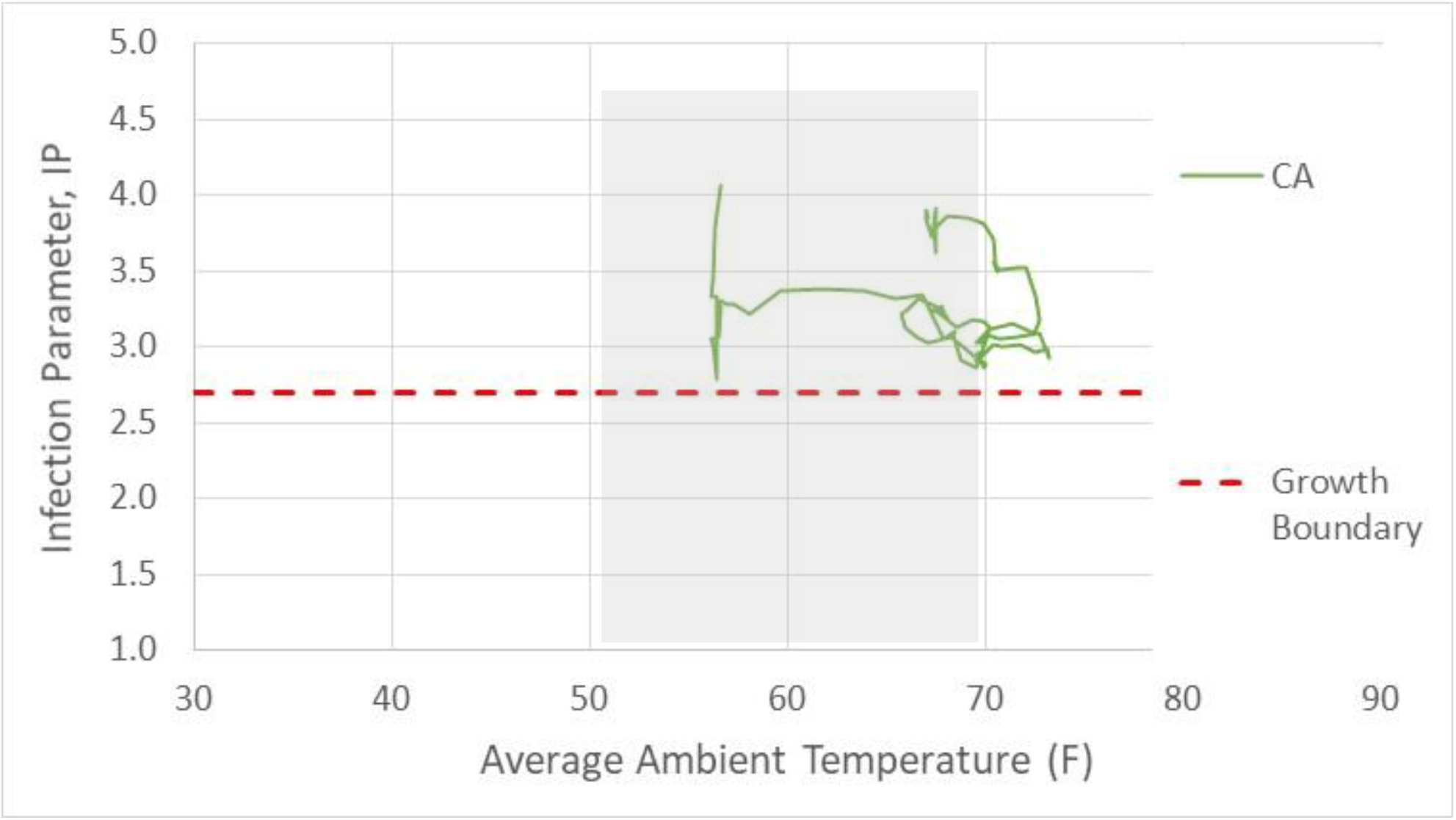
California Infection Parameter trends with outdoor daily average ambient temperature (Los Angeles).

A major metropolitan region was selected to represent each state’s average daily temperature. Figure 19 shows New York state IP data plotted against New York City average daily temperatures over the past 4 months. Note the drop in IP when outdoor ambient temperature reached 50F, followed by a constant IP value between 50F and 70F. The recent abrupt increase in IP is observed to occur with an elevation of outside ambient temperature to 70F.

Figure 20 for Illinois shows a declining IP value at lower temperatures, due to the simultaneous actions of Illinois social distancing efforts and the impact of outdoor ambient temperature increasing above 50F. Chicago, the primary center of infections in Illinois, has been used for temperature data. Illinois reached its minimum IP as outside ambient temperatures reached 65F. Once outdoor ambient temperatures exceeded 70F, similar to New York, an abrupt increase in IP occurred.

Ohio (Figure 21, Columbus used for outdoor temperatures), Michigan (Figure 22, Detroit used for outdoor temperatures), and Minnesota (Figure 23, Minneapolis used for outdoor temperatures) show remarkedly similar trends with a low, constant IP value between 50F and 70F, and abrupt increases in IP for outdoor temperatures below 50F and above 70F when people sealed themselves indoors for heating and cooling comfort, respectively.

California (Figure 24, Los Angeles) is geographically and climatically diverse, and representing such a large area with one city is unreasonable. Initial Covid-19 outbreaks occurred in the Bay area, however, these outbreaks were contained as travel and social distancing efforts were enacted. Since the early outbreaks, southern California has been the primary center of infection activity. Although more poorly defined, Figure 24 shows trends similar to northern states with a relatively low IP at temperatures below 70F, and elevated IP as outdoor ambient temperature in Los Angeles increased above 70F.

Is it a coincidence that outdoor temperatures from 50F to 70F, are coincident with the lowest levels of Infection Parameter? Similarly, is it a coincidence that an abrupt, rather finite jump in infection parameter occurs when homes are being sealed up and closed for heating and cooling operation?

## Reduction of Indoor Infection Probability

The reduction of infection parameter during “nice” weather is not a coincidence. Buildings in the US, residential, commercial, institutional, religious and others are poorly ventilated and have poor indoor air quality. Improving ventilation in buildings will decrease Covid-19 disease transmission.

Figure 8 gives a sense of the range of improvement. If there is no difference between the indoor environment and outdoor environment, we should not have observed an abrupt change in infection parameter and disease transmission efficiency parameter as outdoor temperatures drove people into buildings. Without any change in social distancing (SDI=33), and no change to indoor environments, we follow the upper path and will see daily infections rise well above 100,000 infections per day. It is likely that people will continue distancing as infection numbers continue to increase, perhaps to SDI levels above 36 where the infection growth/decay boundary (IP=2.7) occurs, causing a reduction in infection cases as shown by the intermediate path in Figure 8.

The impact of changes to an indoor environment’s ventilation system can be assessed using the Wells-Riley disease transmission modeling of airborne diseases (3). We note that one does not need to become muddled in the debate between airborne transmission and other modes (direct and fomite). Ventilation impacts all disease transmission modes. Poor ventilation creates densely covered infectious surfaces (fomites), and poorly ventilated buildings extend the range, circulation, and airborne duration of so-called “big droplets” (5 microns and larger) that take several minutes to “fall” to the floor.

Monitoring carbon dioxide concentration is key to understanding the indoor environment. Carbon dioxide concentration should be reduced below 800ppm, which indicates fresh air ventilation and infiltration levels that are double today’s odor-based ventilation standards. Milton and colleagues (4) found this level of ventilation air flow reduced the incidence of sick days by 40% (equivalent to seasonal flu vaccine effectiveness) in a study of over 3000 employees spread among several buildings and work environments. Flu and cold infections are respiratory illnesses that are not considered to be airborne, similar to SARS-CoV-2. Regardless of the dominant transmission mode, ventilation is extremely important.

Following Rudnick and Miltons’ work (3), Figure 25 relates indoor carbon dioxide concentration levels to infection probability and exposure time. A virus shedding rate of 100quanta per hour per infectious person is assumed. Figure 25 also assumes 3 susceptible and 1 infectious occupants in the building.

**Figure 25.**
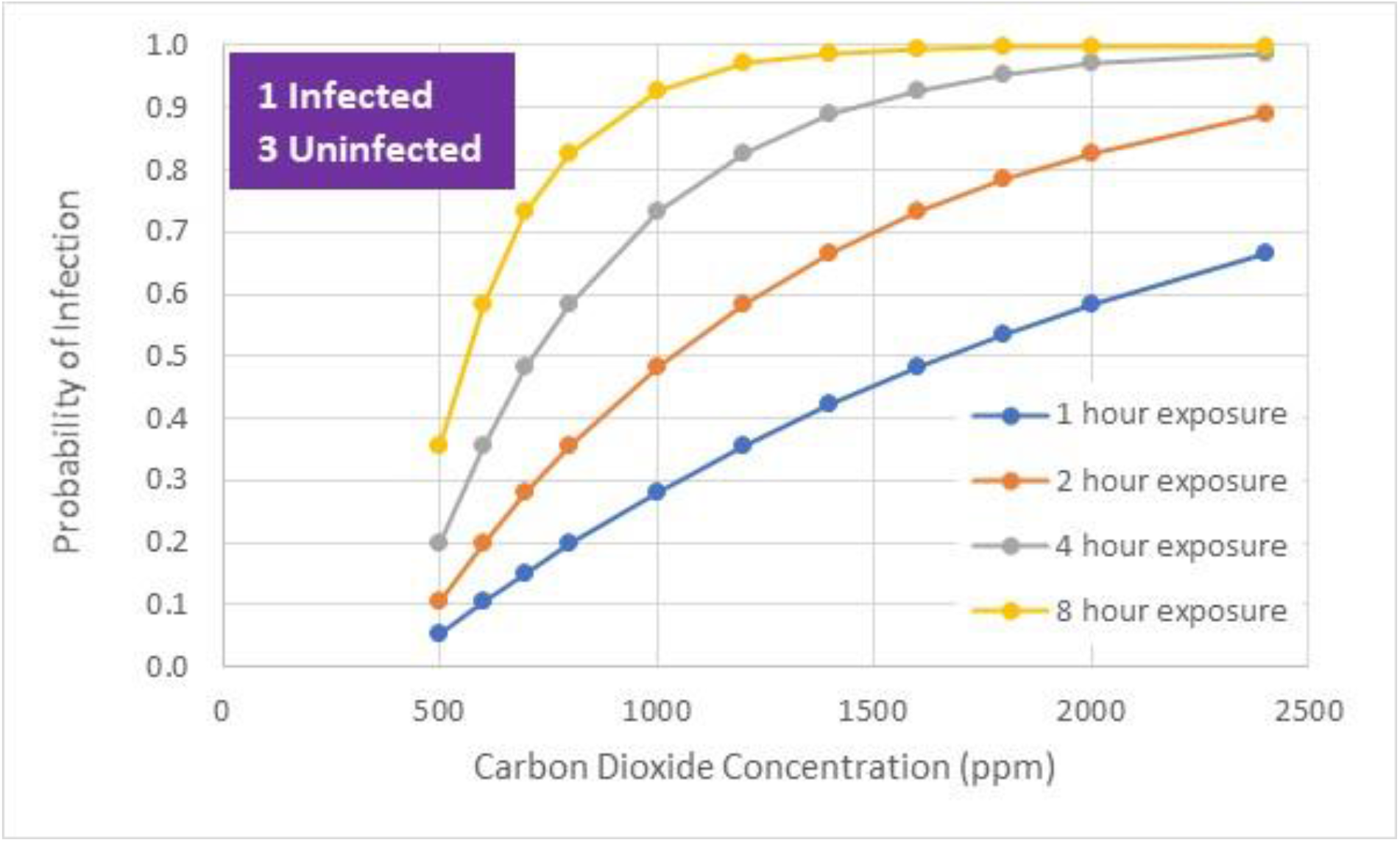
Infection probability as a function of carbon dioxide concentration and exposure time.

Figure 26 shows three residential ventilation standards (ASHRAE 62.2-2010, ASHRAE 62.2-2019, and Passive House 0.3 air changes per hour), the common “rule-of-thumb” ventilation level of 20cfm per person, and enhanced ventilation of 40cfm per person. Building occupants are assumed to wear masks that reduce quanta shedding by 50%. All three standard residential ventilation standards result in 90% probability of infection over an 8 hour exposure period. A building with 40cfm, improved air filtration (MERV 13), and UVGI (0.2 Wuv per cfm air flow) reduces infection probability to 30%.

**Figure 26.**
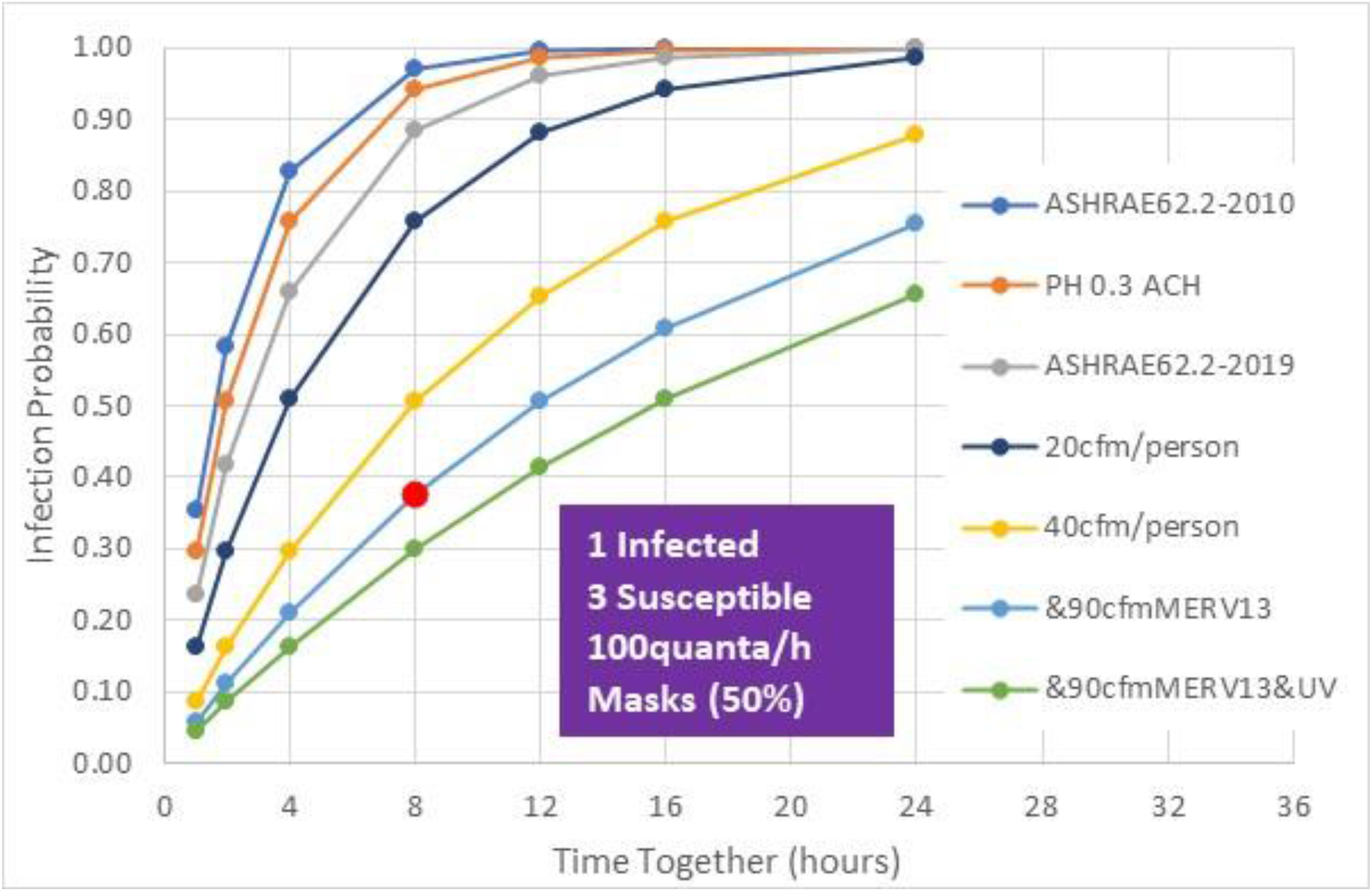
Reduction of Covid-19 infection probability with increased fresh air, improved air filtration, UVGI and face masks.

Relating the impact of enhanced ventilation guidelines to the transmission efficiency parameter is difficult without collection of data, however, the significant difference between standard building practice and enhanced ventilation guidelines indicates a significant impact to disease transmission when weather conditions outside the 50F to 70F range force people indoors.

## Summary

Ironically, today’s building comfort conditioning equipment is causing the opposite effect with the SARS-CoV-2 pandemic in contrast to the lack of summertime air conditioning technology during the 1918 pandemic. Covid-19 transmission efficiency is nearly doubled in relation transmission efficiency in the 50F to 70F swing season window. Increased fresh air flow, improved air filtration and installation of UVGI can significantly reduce infection probability within buildings. These improvements will also result in improved cognitive performance (5,6) and reduced sick days that have significantly more value than energy costs associated with increased ventilation rates (6).

A simple, cost-effective manner for determining air quality in homes and building spaces is carbon dioxide monitoring. Building occupants in schools, office buildings, shops, restaurants, and other public gathering spaces should carry carbon dioxide monitors to their work place, school or other public buildings with unknown ventilation levels and occupancy schedules. A $200 carbon dioxide sensor with 30ppm accuracy is sufficient. Indoor carbon dioxide should be kept below 800ppm, which is equivalent to doubling today’s fresh air ventilation standards. The impact on Covid-19 transmission will require field data and analyses to determine, however, these enhancements to building ventilation systems will improve cognition and reduce sick days that are worth much more than the cost of additional ventilation energy.

## Data Availability

All data used in the analyses are cited and available

https://www.worldometers.info/coronavirus/

https://data.covid.umd.edu/

http://91-divoc.com/

https://www.wunderground.com/

## Appendix A – Covid-19 Infection Growth Model

An early-stage infection growth model is used to demonstrate infection growth based on social distancing. The model assumes infection growth rate to be proportional to the number of infectious cases in a fully susceptible populace:

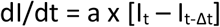

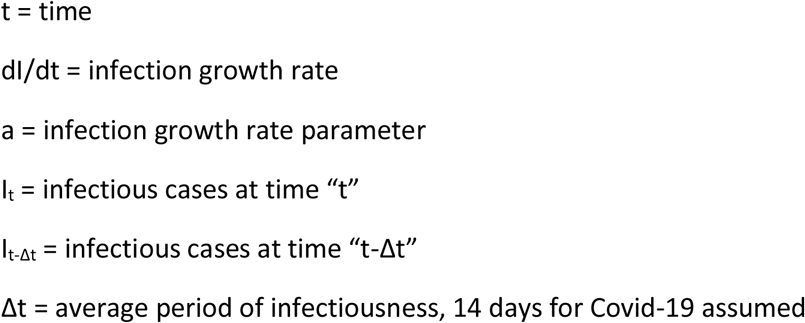

Future infections can be predicted with a forward time step algorithm.

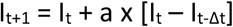

The infection growth parameter can be determined from empirical data as:

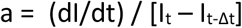

A daily growth parameter can be numerically computed as:

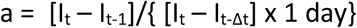

## Appendix B - Infection Parameter, IP

The growth parameter, a, can be scaled to produce a more descriptive parameter for understanding infection growth. We define the infection parameter, IP, as the ratio of current infections to infections two weeks prior:

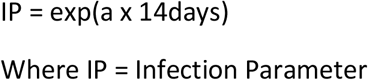

IP ranges from 1 to much greater than 1, as shown in Figures B1 and B2 for 8 countries. For example, for a daily growth factor of 0.1 per day, IP is 4. During the latter part of March, caused by delayed social distancing restrictions, IP values in the US soared to greater than 50, primarily driven by uncontrolled infection growth in New York. By April 7, New York and other states (eg, Michigan, Illinois, New Jersey) with high infection spread were able to implement restrictions that dropped IP below 7. Unfortunately, “distancing fatigue” in the US is preventing IP from dropping below 2, where Covid-19 recoveries could outpace new infections.

**Figure B1.**
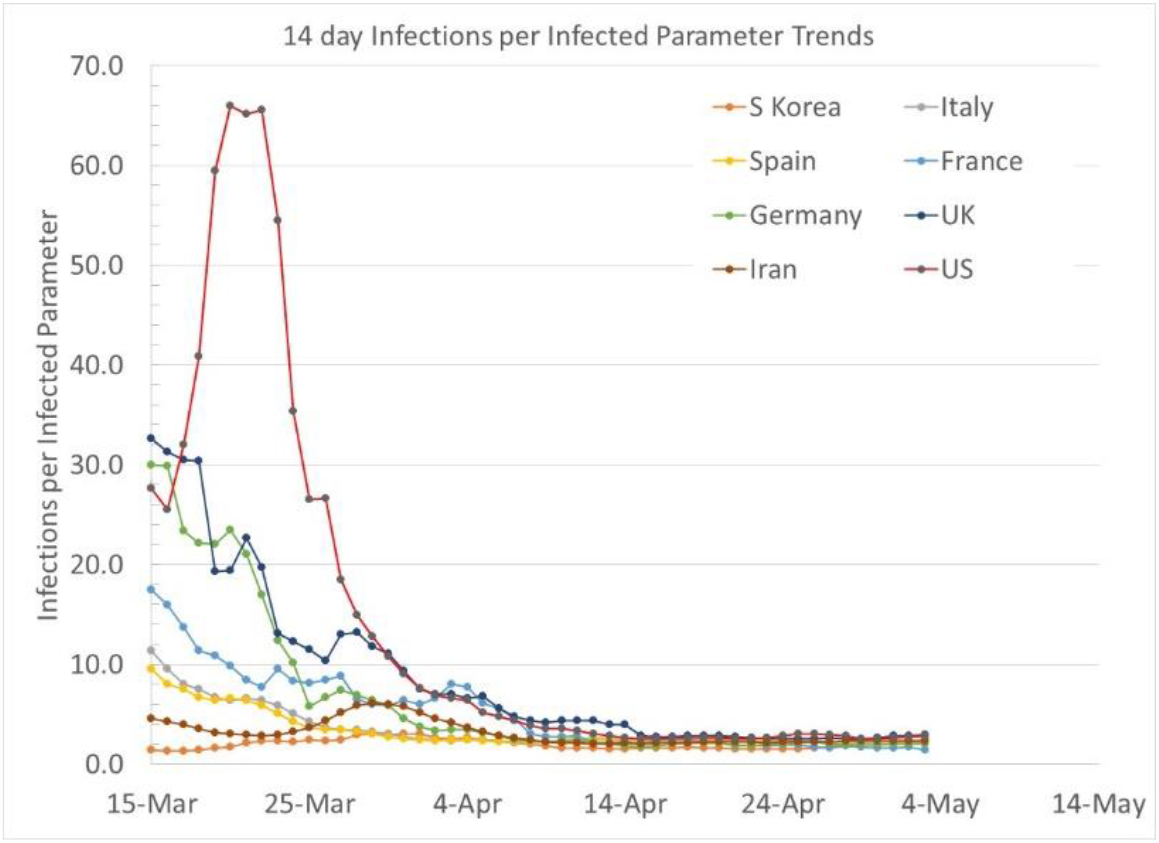
Infection Parameter, IP, trends for 8 countries since March 15, 2020

**Figure B2.**
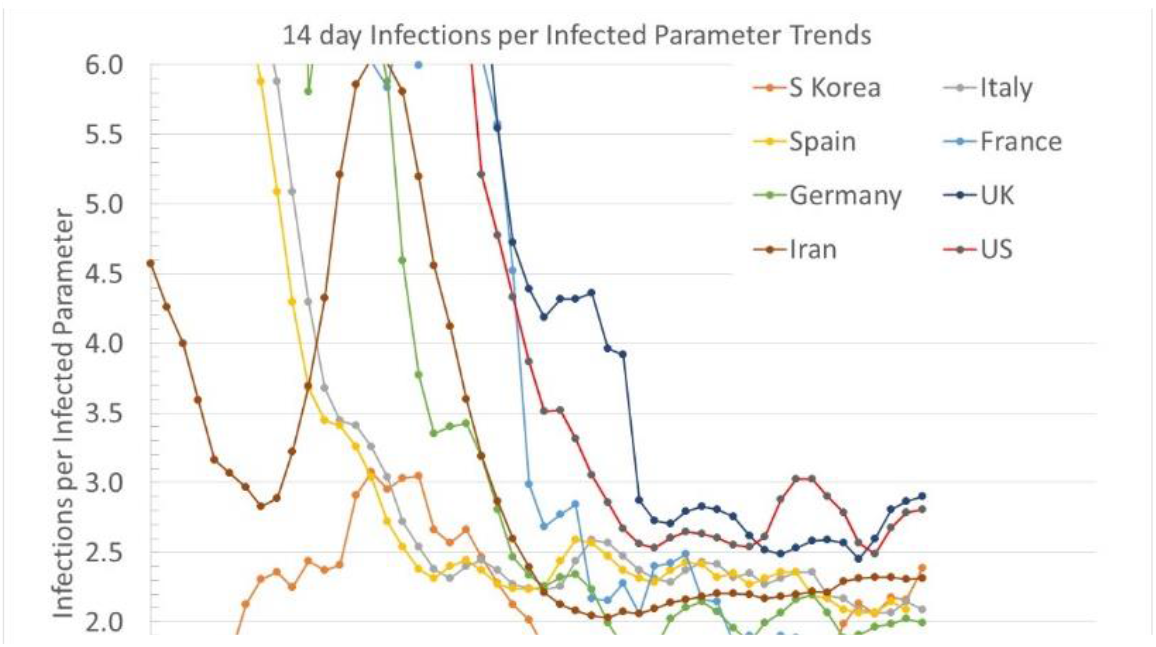
Infection Parameter, IP, trends at lower IP values for 8 countries.

IP qualitatively describes the number of infections incurred over a 14 day period per infectious person. IP is related to the Basic Reproduction number, R_0_, and is a function of factors related to disease transmission efficiency, social contacts, asymptomatic carriers, seasonal/climatic effects, indoor environmental factors, age, face masks usage, building ventilation, and other conditions.

Linear infection growth is a boundary between new infection growth and decay. One can show that linear infection growth results in an Infection Parameter (IP) value of “e” (∼2.72). When IP is greater than “e”, daily new infections are growing and when IP is less than “e”, daily infections are decaying. At an IP of “e”, daily new infections are constant.

## Appendix C – Correlation of Infection Parameter and Social Distancing Index

The Infection Parameter can be related to two, assumed independent factors, a social distancing index (SDI) and a disease transmission efficiency parameter (G). For example, people may restrict their movements (increased social distancing) but continue to shake hands, hug, and not wear face masks. Alternatively, social distancing may be decreased as businesses and social activities re-open, but disease “transmission efficiency” can be reduced by people not hugging and by wearing face masks. Other factors such as building ventilation characteristics (fresh air flow per occupant, air filtration and sanitation) also impact the transmission efficiency. We relate the Infection Parameter to a social distance index in this section, and discuss transmission efficiency in appendix D.

The University of Maryland social distancing index (SDI) is a parameter based on anonymous cell phone and vehicle gps data (reference 1C). The SDI incorporates weighted factors describing social interactions, and is scaled between 0 (maximum social interaction) and 100 (minimum social interaction). “Normal” weekday SDI values, prior to social distancing in the US, were somewhat less than 20. Social distancing restrictions implemented in the US around the 3^rd^ week of March, increased weekday SDI to 50 with weekends reaching 70.

Relating SDI to IP consists of two operations:

a. Computing a 14 day moving average of daily SDI values, as shown in Figure C1
b. Correlating IP and SDI with a 7 day phase shift, as shown in Figure C2

The 7 day phase shift correlates a daily IP value with the moving average SDI value 7 days prior to the IP value (ie, the IP value before March 21 paired with the 14 day moving average value of SDI from March 14). The phase shift between today’s IP value and the SDI value from 7 days previous accounts for incubation of the disease. The surprisingly strong correlation between IP and SDI based on the moving average and phase shift provides a simple predictive model of Covid-19 transmission:

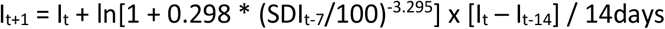

State SDI values are also available from the University of Maryland database. 14 day moving average SDI values for a number of states are shown in Figure C3, with trends similar to the US SDI, however, note different levels of SDI as well as variations of transient effects in SDI variations. For example, New York reached the highest level of SDI, and has continued with a high SDI level. Florida is showing the impact of recent Covid-19 case surge with increasing SDI levels in comparison to other states with less severe infection spread.

---

**Figure C1.**
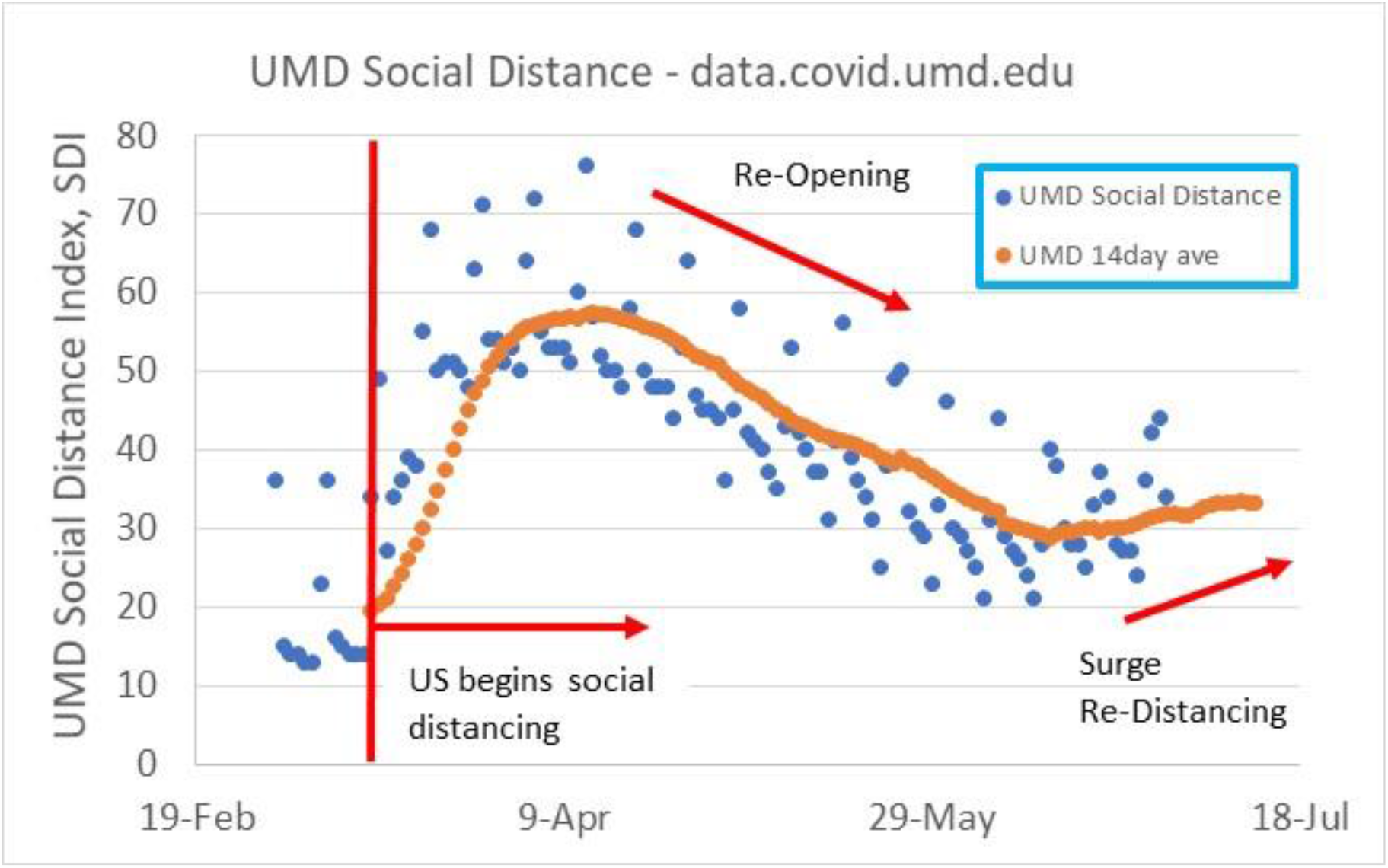
Daily and 14 day moving average UMD social distancing index values.

**Figure C2.**
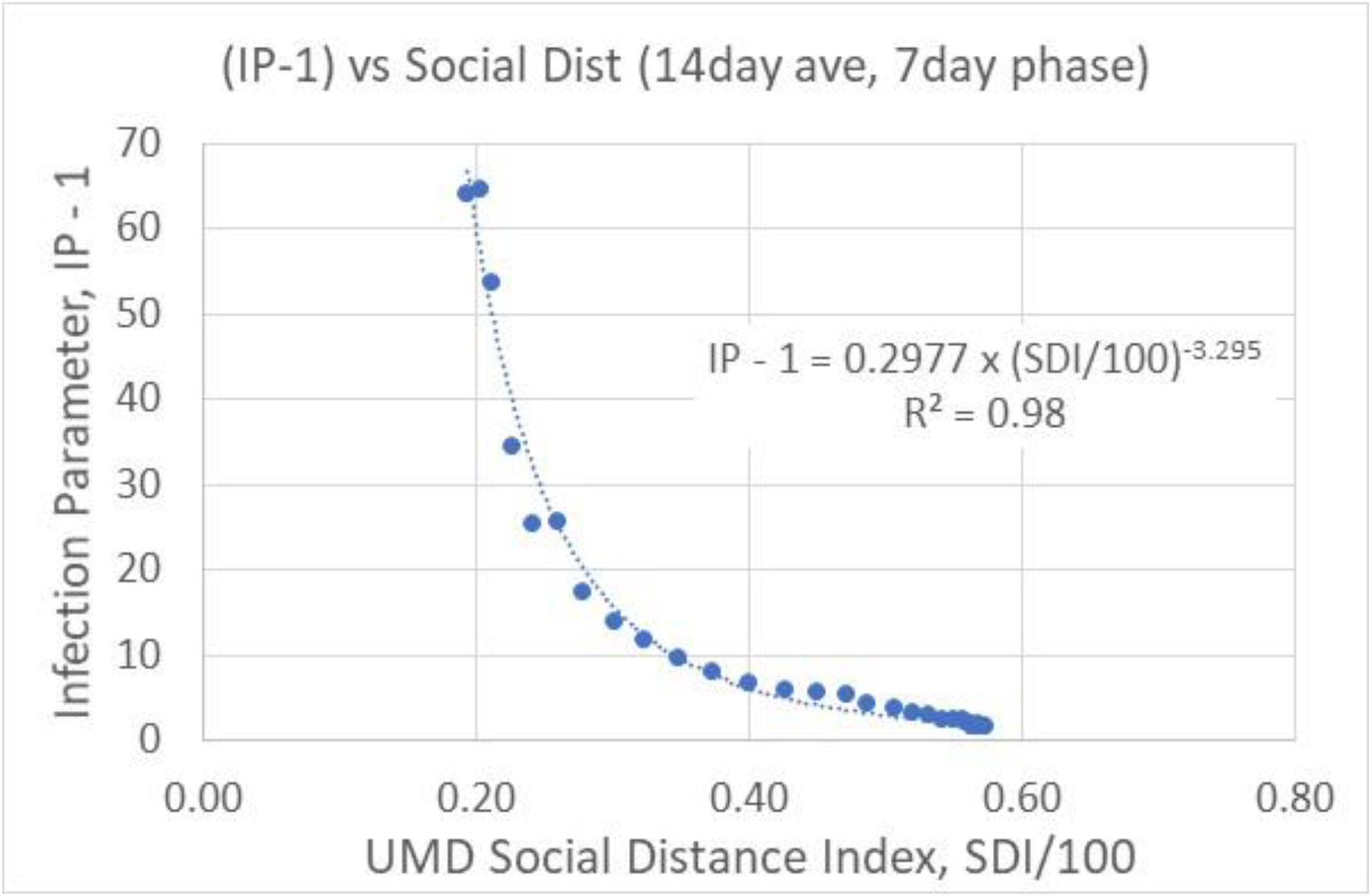
Correlation of Infection Parameter, IP, and UMD social distance index, SDI.

**Figure C3.**
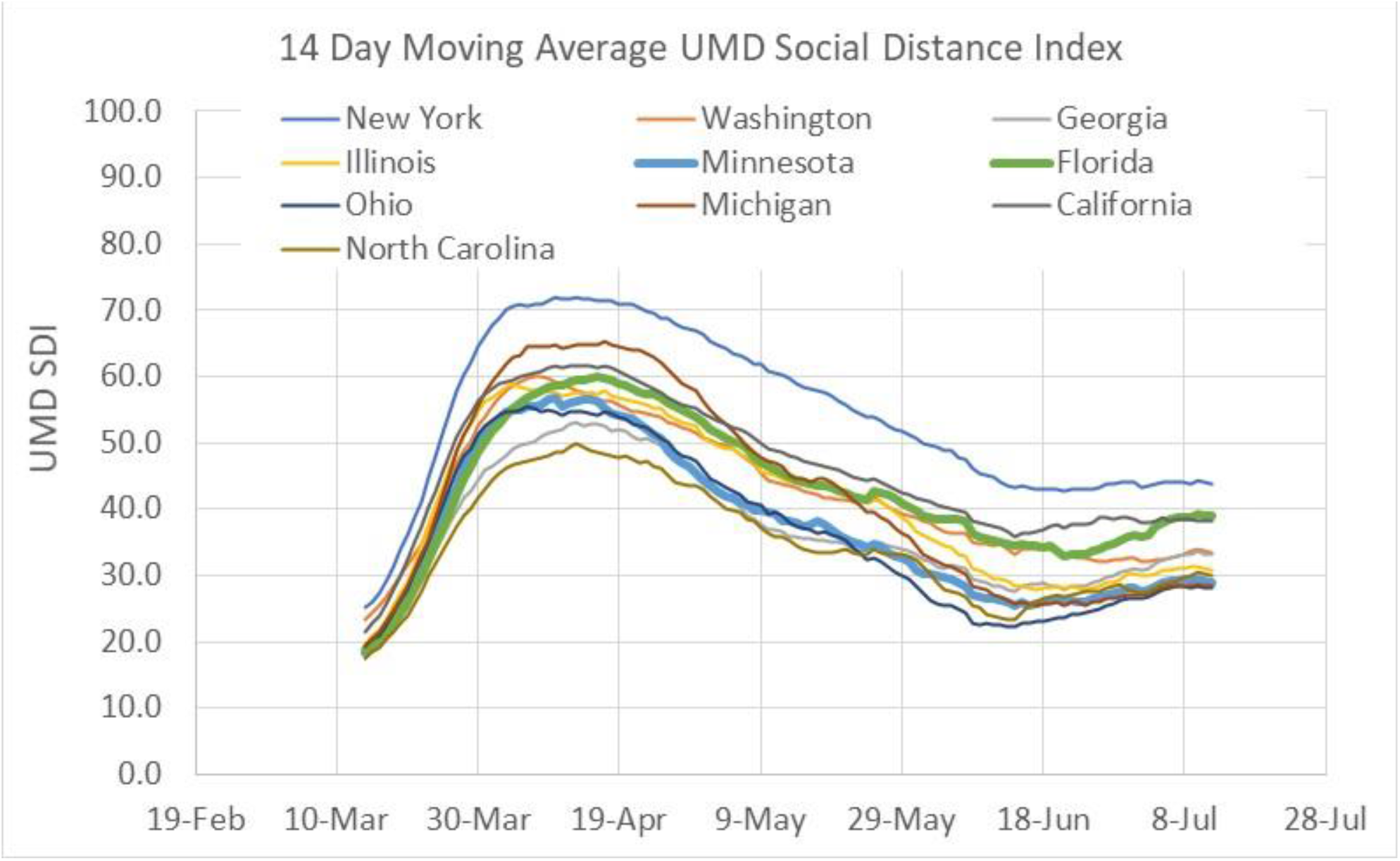
State SDI trends over time (14 day moving average SDI)

## Appendix D SARS-CoV-2 Disease Transmission Efficiency

A disease transmission efficiency parameter, G, is defined as the ratio of Infection Parameter at a given Social Distance Index to the IP that occurred during the early stages of social distancing in the US prior to implementation of significant control measures, such as face masks and 6 ft distancing. The time period between early March 2020 and early April 2020 (see Figure C1) with increasing social distancing is used to define this time period. Late April and beyond in the US is characterized as a time when face masks, 6 ft distancing, no hand shaking, no hugging, and other methods for decreasing disease transmission efficiency were implemented. Transmission efficiency can be greater or less than 1.

Figure D1 shows the trend in US IP values versus SDI for the early March-April 2020 correlation used to define a disease transmission efficiency of 1. Parametric disease transmission efficiency lines for G equal to 0.6, 0.3, and 0.1 are drawn on Figure D1. “IP new” values plotted on Figure D1 are data since April 2020 as disease transmission reduction efforts were enacted. As the US “re-opened”, SDI values were reduced to 30 coupled with lowered disease transmission efficiency.

Figures D2 through D10 show IP versus SDI and G trends for several states (New York, Washington, Georgia, Illinois, Florida, Ohio, Michigan, California and North Carolina). Note initial social distancing trends in which some states, such as New York, had initial disease transmission efficiency levels above the average US level used to define G=1. Some states, such as Washington, displayed disease transmission efficiency levels lower than 1 during initial social distancing efforts.

Figures D11 and D12 show the loci of SDI values versus initial disease transmission efficiency levels (ie, “normal” social interactions before face masks, 6 ft distancing, etc) that result in an IP level of “e”, the boundary between infection growth and decay. Figure D11 shows that the US must have an SDI isolation of 60 with normal interactions in order to avoid growth of Covid-19 infections. Figure D12 shows SDI isolation levels for individual states based on their initial disease transmission characteristics. New York and Illinois, due to high disease transmission efficiency in New York City and Chicago disease epicenters needed to reach SDI levels above 65, while Washington had a relatively low disease transmission efficiency and required an SDI of 50 in order to reduce infection growth.

**Figure D1.**
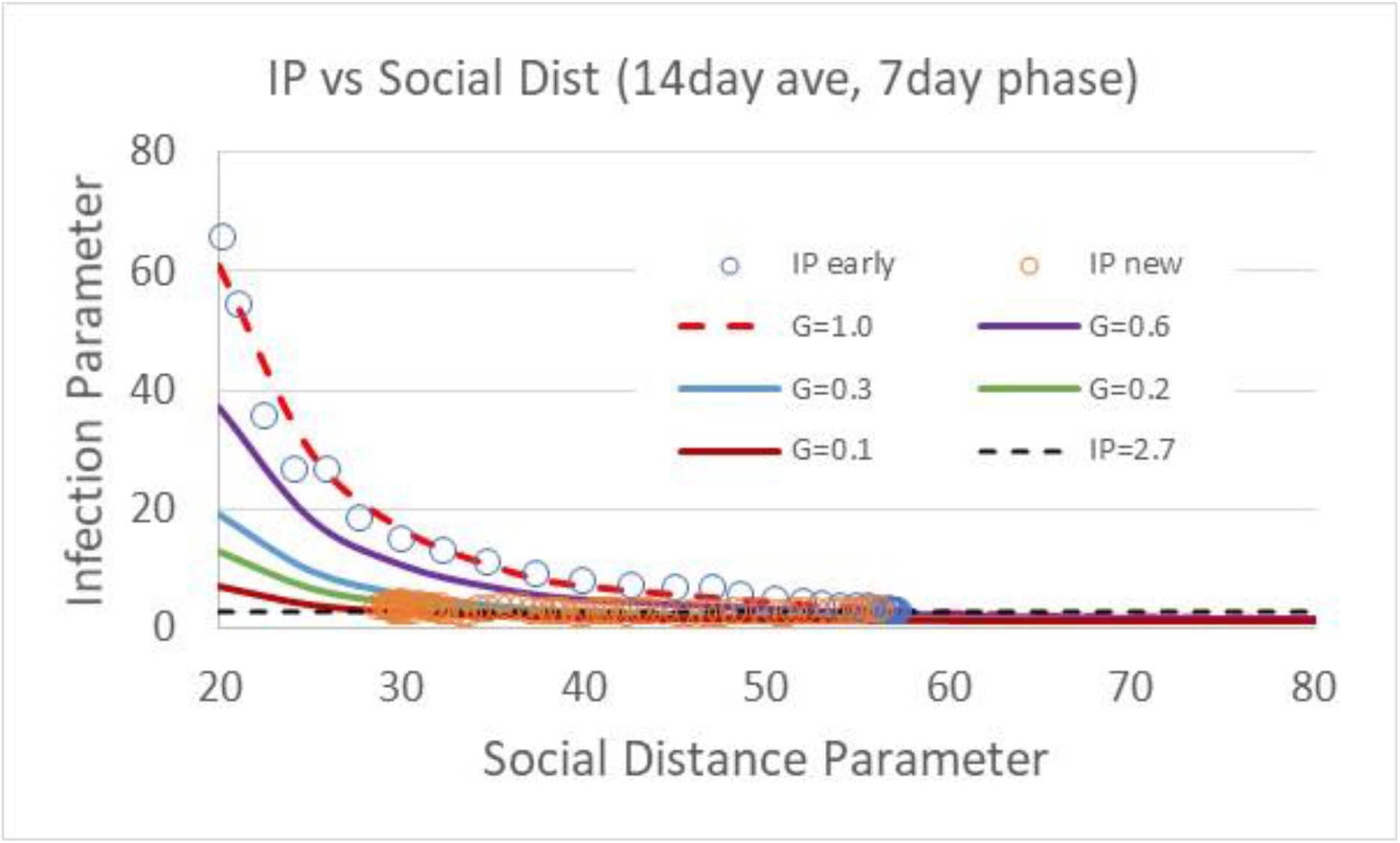
IP versus US SDI with parametric lines of transmission efficiency, G. plotted. Blue open circule data are early (March and April 2020) data during US shutdown used to define transmission efficiency of 1.

**Figure D2.**
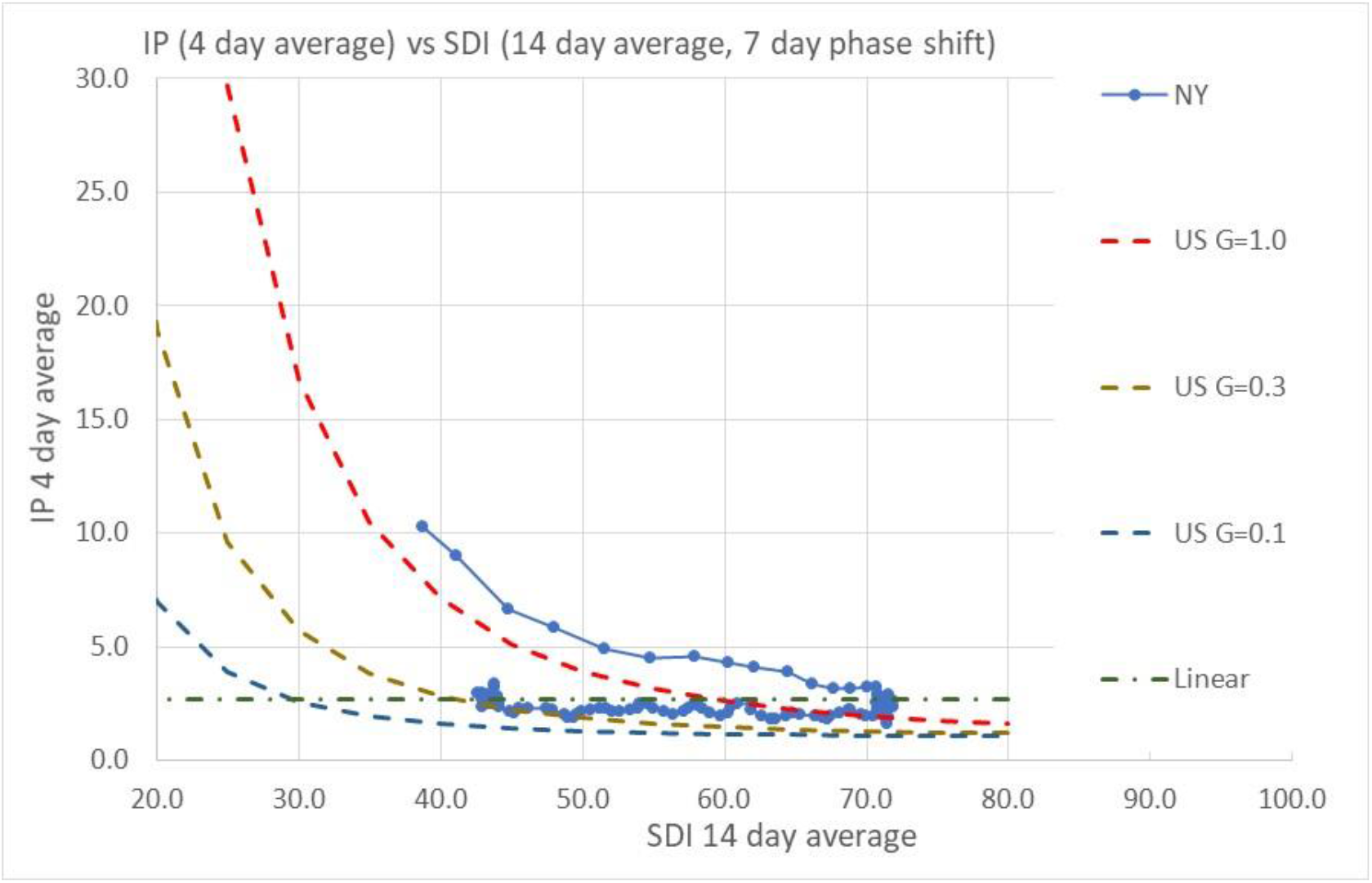
New York IP trends as a function of SDI and G. Initially, NY IP has a transmission efficiency greater than 1, perhaps reflecting closer interactions among people in NYC due to denser housing, public transportation, and other factors.

**Figure D3.**
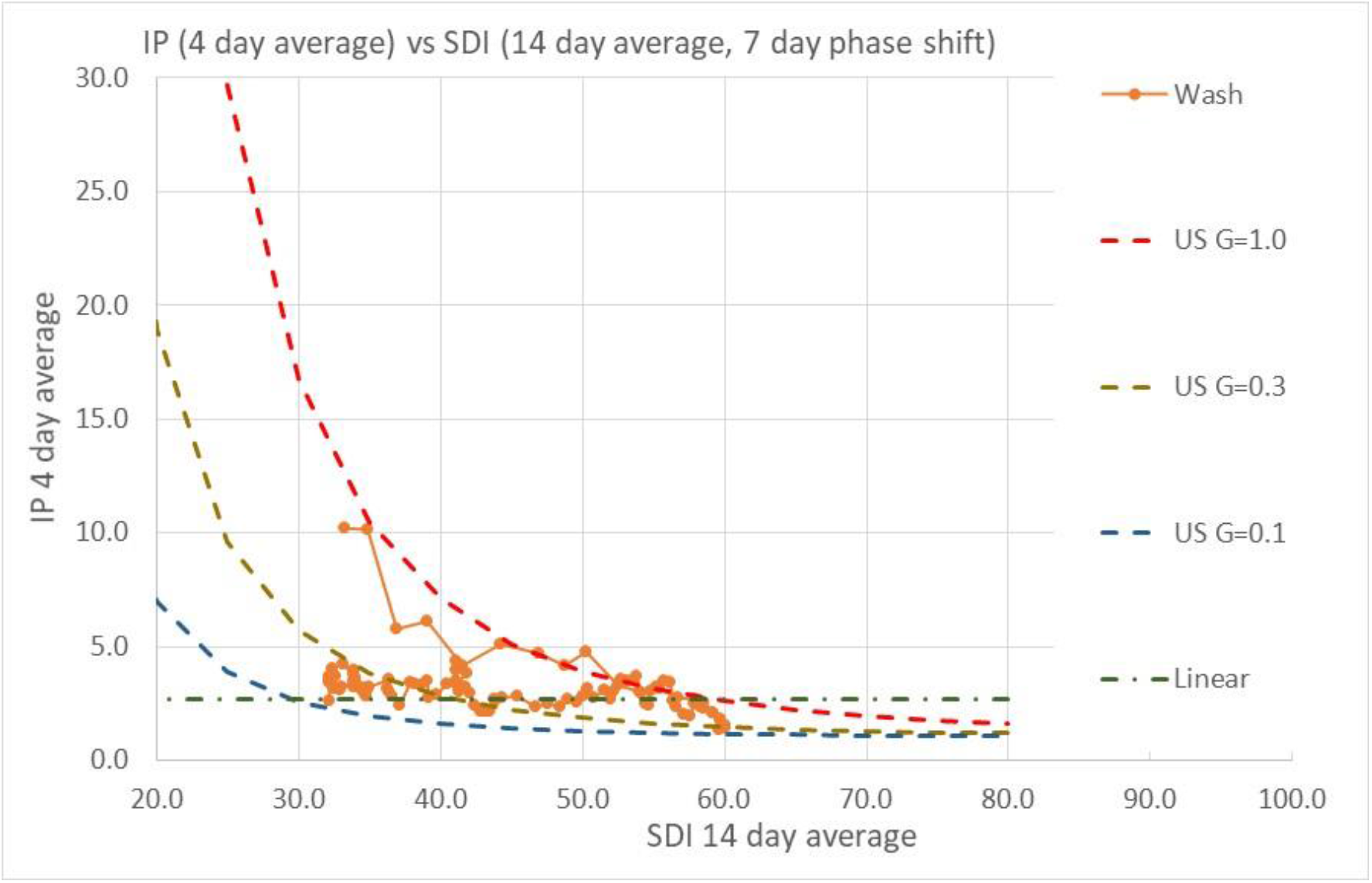
Washington IP trends as a function of SDI and G.

**Figure D4.**
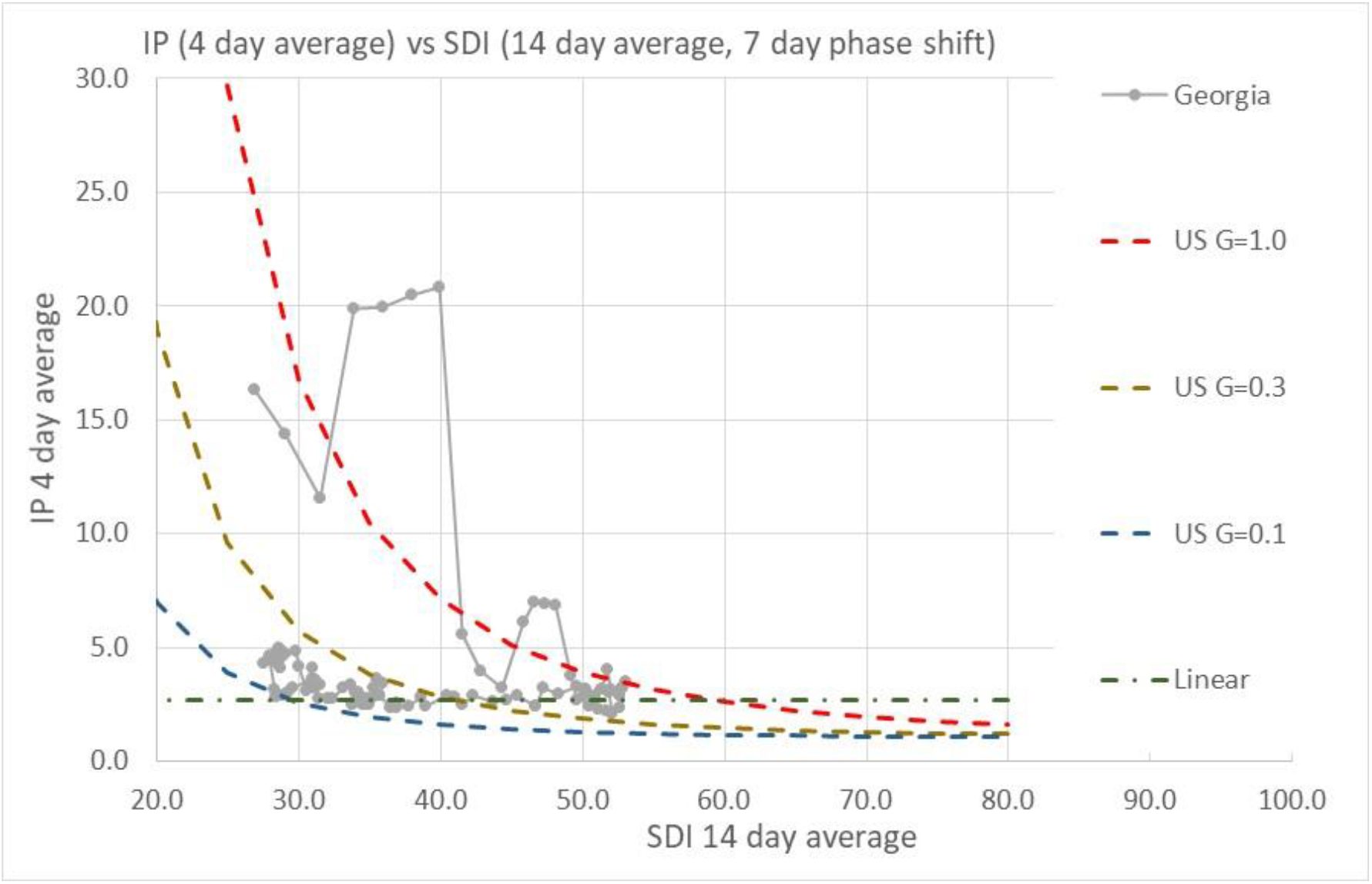
Georgia IP trends as a function of SDI and G.

**Figure D5.**
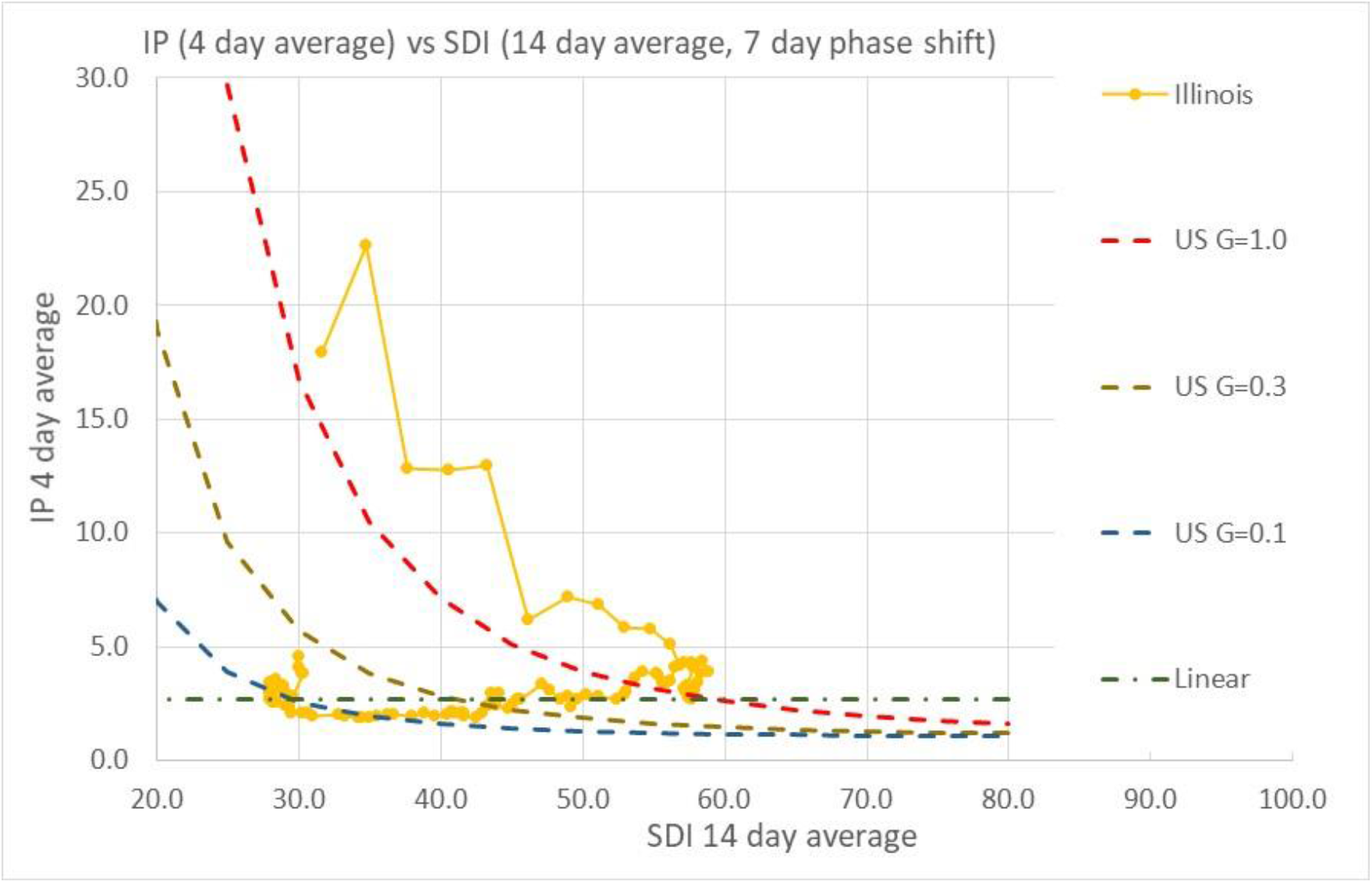
Illinois IP trends as a function of SDI and G.

**Figure D6.**
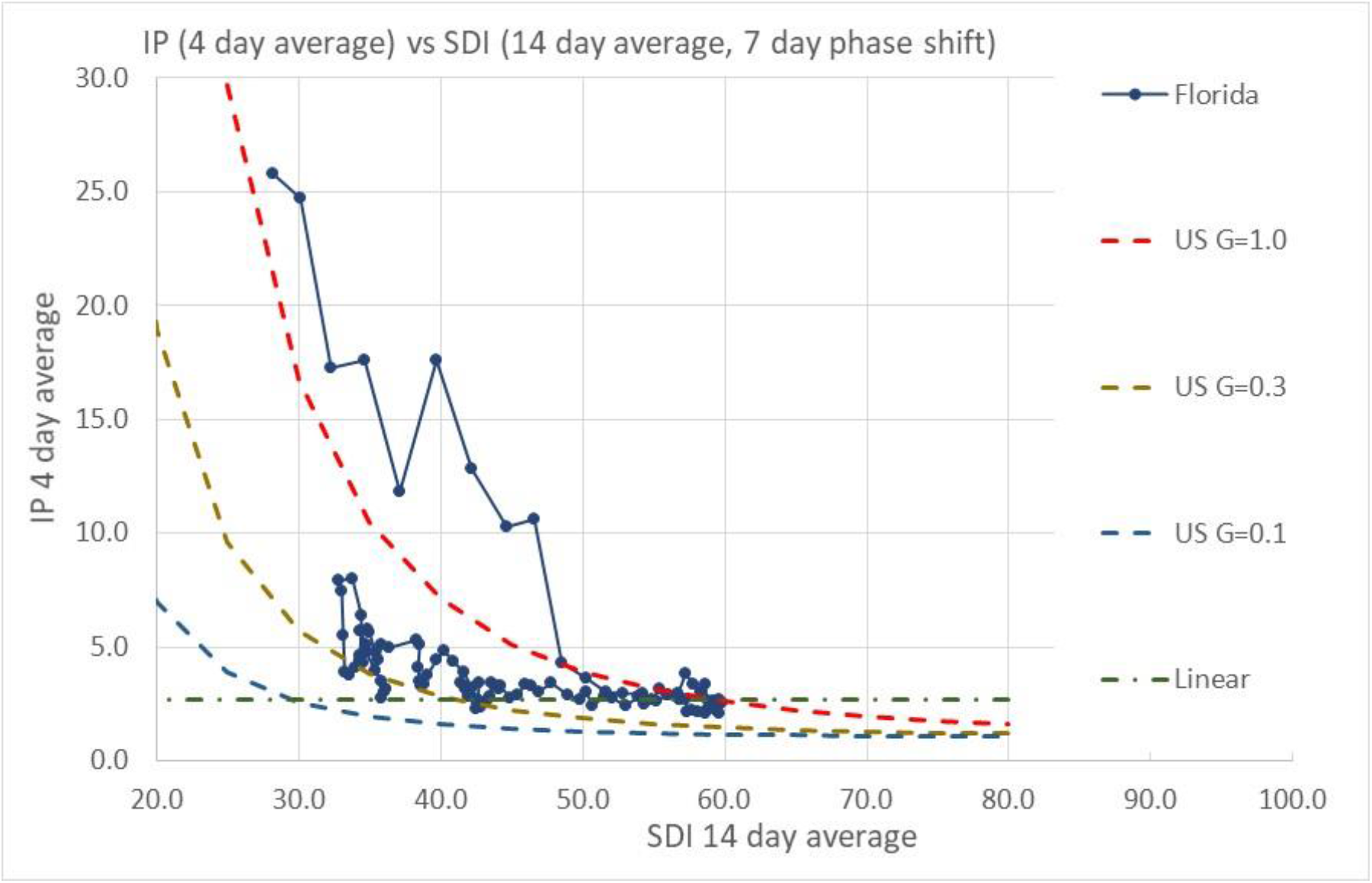
Florida IP trends as a function of SDI and G.

**Figure D7.**
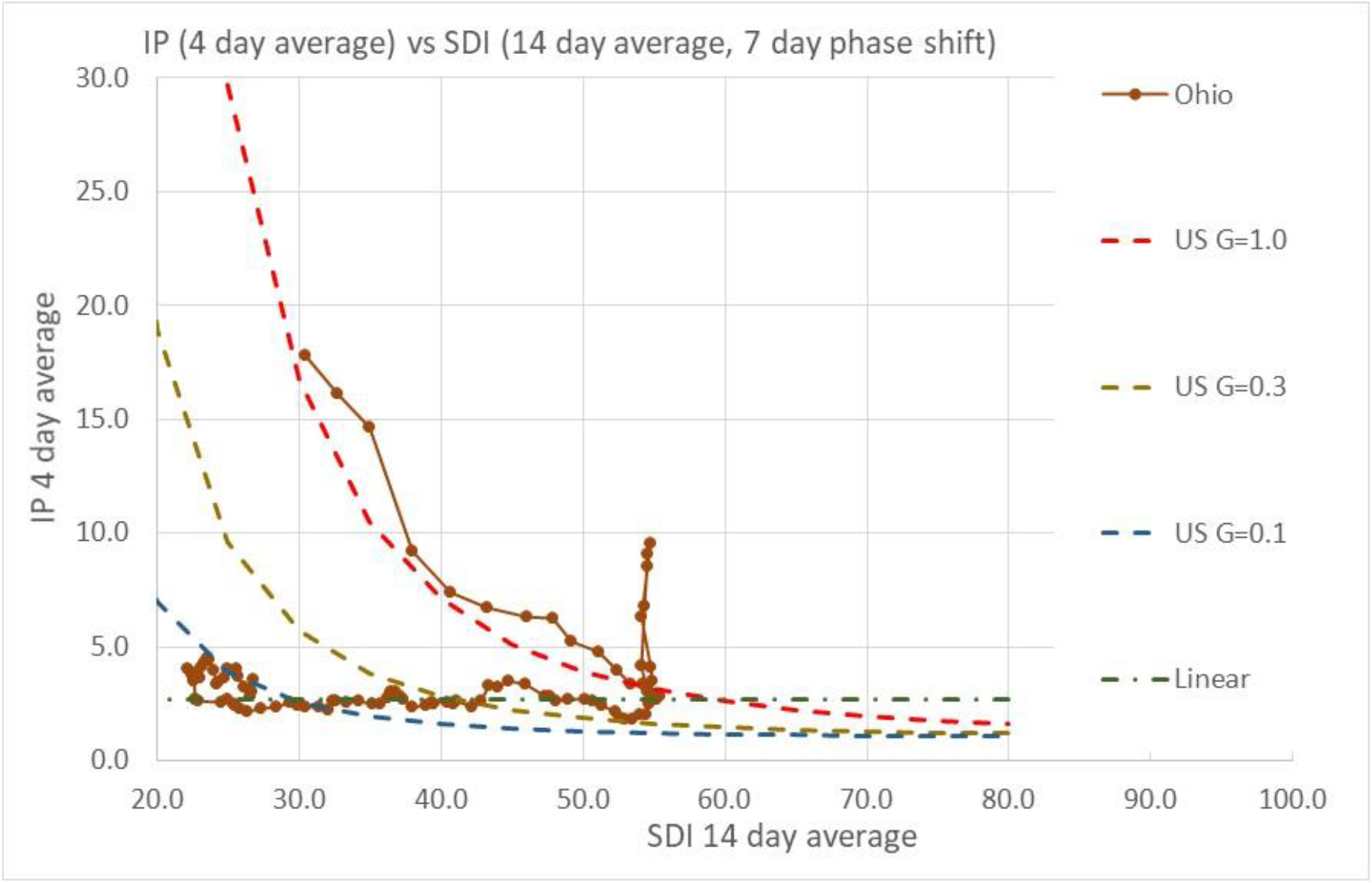
Ohio IP trends as a function of SDI and G. Note the infection “spike” during a localized infection outbreak in a prison.

**Figure D8.**
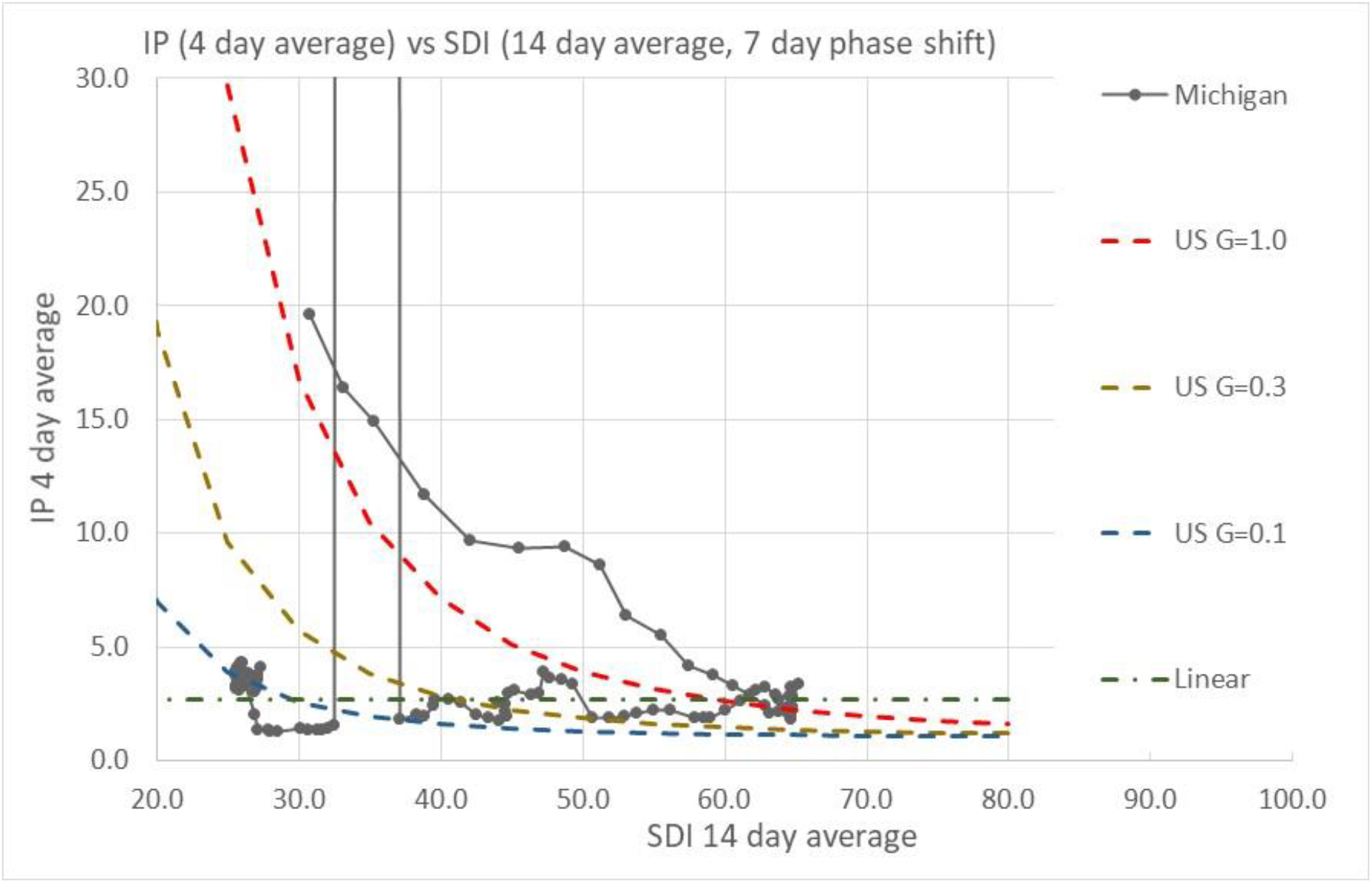
Michigan IP trends as a function of SDI and G. The jump in IP values is due to a change in data reporting.

**Figure D9.**
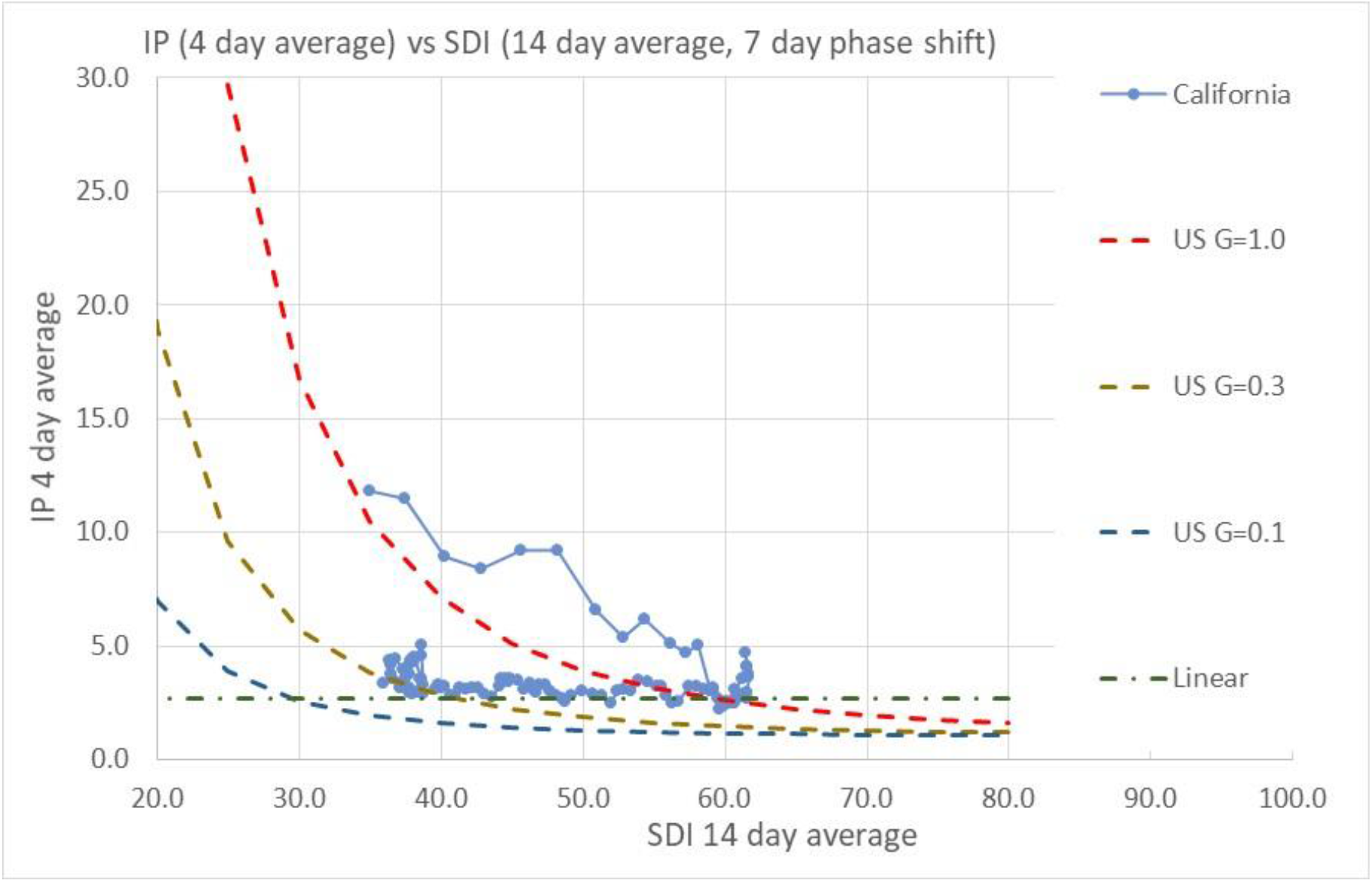
California IP trends as a function of SDI and G.

**Figure D10.**
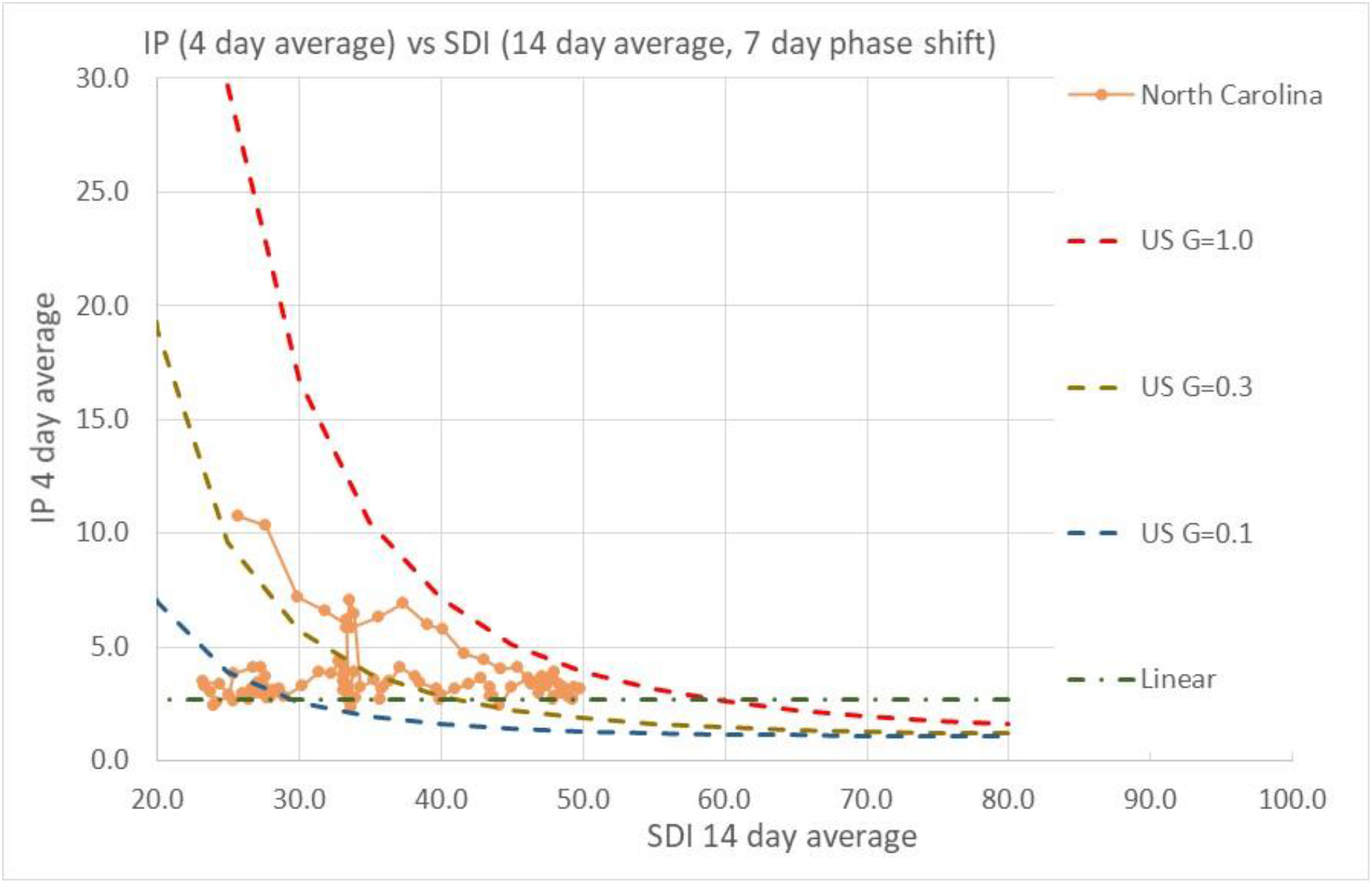
North Carolina IP trends as a function of SDI and G.

**Figure D11.**
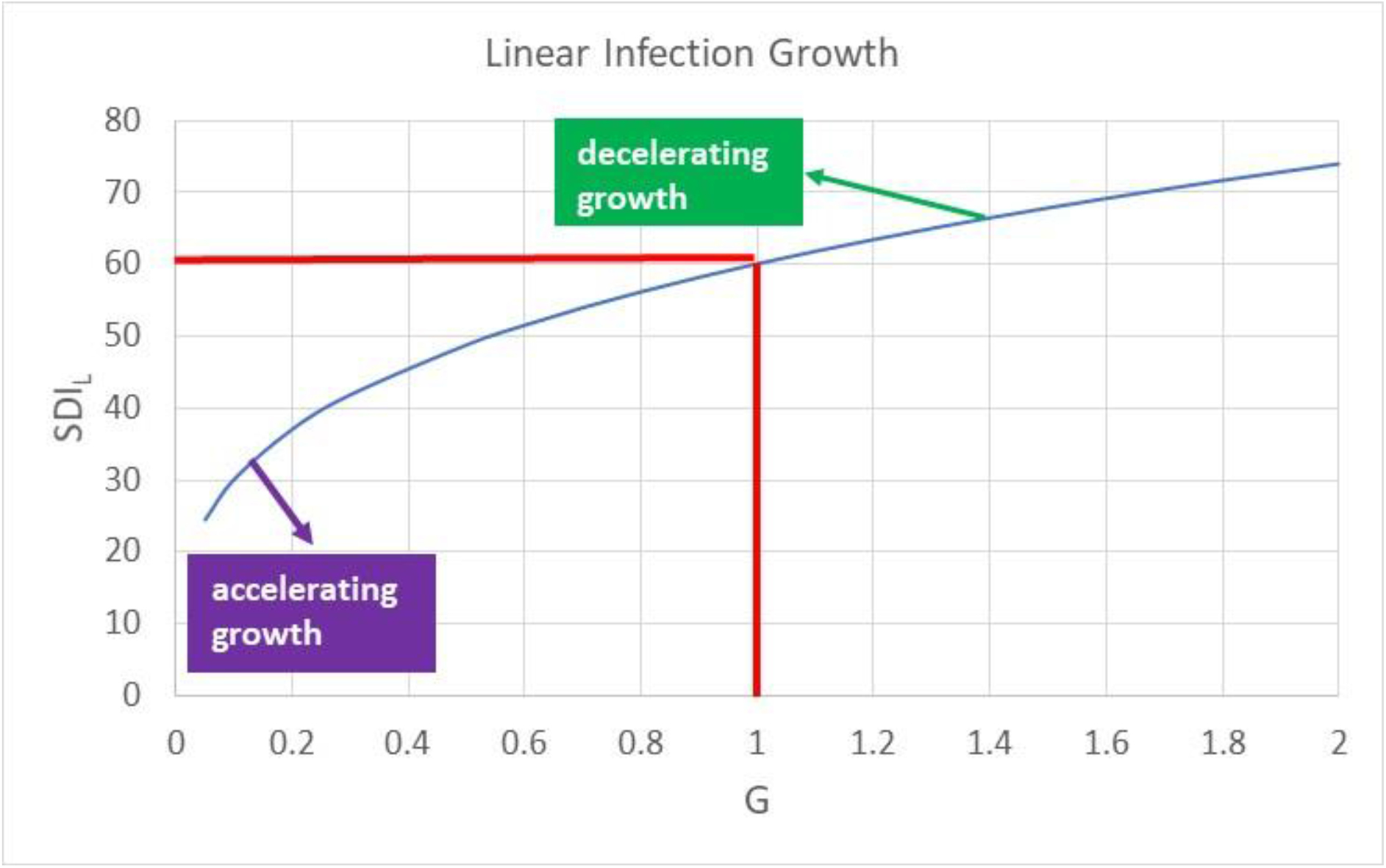
Relation between SDI and G defining region of growing daily infections and decreasing daily infections.

**Figure D12.**
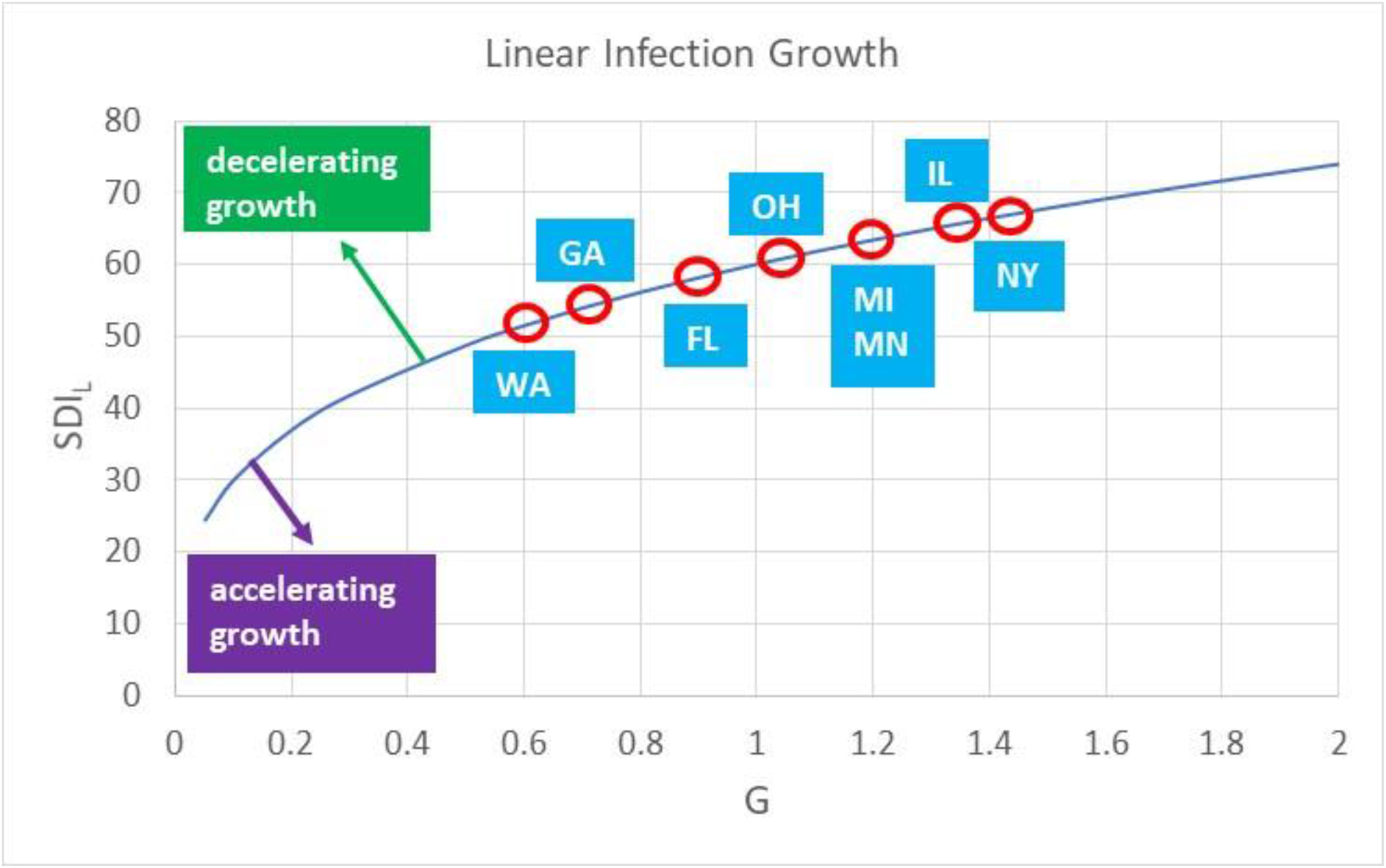
SDI levels required for individual states based on initial levels of G showing SDI required to avoid daily growth of infections.

